# BayesSMILES: Bayesian Segmentation Modeling for Longitudinal Epidemiological Studies

**DOI:** 10.1101/2020.10.06.20208132

**Authors:** Shuang Jiang, Quan Zhou, Xiaowei Zhan, Qiwei Li

**Affiliations:** Department of Statistical Science, Southern Methodist University, Dallas, TX 75205, USA; Quantitative Biomedical Research Center, Department of Population and Data Sciences, The University of Texas Southwestern Medical Center, Dallas, TX 75390, USA; Department of Statistics, Texas A&M University, College Station, TX 77843, USA; Department of Mathematical Sciences, The University of Texas at Dallas, Richardson, TX 75080, USA

**Keywords:** Bayesian hierarchical modeling, Multiple change-point detection, Poisson segmented regression, Stochastic SIR model

## Abstract

The coronavirus disease of 2019 (COVID-19) is a pandemic. To characterize its disease transmissibility, we propose a Bayesian change point detection model using daily actively infectious cases. Our model builds on a Bayesian Poisson segmented regression model that can 1) capture the epidemiological dynamics under the changing conditions caused by external or internal factors; 2) provide uncertainty estimates of both the number and locations of change points; and 3) adjust any explanatory time-varying covariates. Our model can be used to evaluate public health interventions, identify latent events associated with spreading rates, and yield better short-term forecasts.

## 1 Introduction

A newly identified coronavirus, SARS-CoV-2, is a lethal virus for humans. It has caused a worldwide pandemic for the disease known as COVID-19. As reported by the Johns Hopkins University Center for Systems Science and Engineering (JHU-CSSE), the COVID-19 pandemic has spread to 188 countries and territories, with more than 14 million confirmed cases by the end of July 2020. The extremely rapid spreading of the disease and the increasing burden on healthcare systems have became major public health problems. In response to the public health demand to “flatten the curve” (Akiyama et al., 2020), both federal and local governments in the United States (U.S.) have enforced a wide range of public health measures, such as border shutdowns, travel restrictions, and quarantine.

As a consequence, the importance of understanding the COVID-19 dynamics is steadily increasing in the contemporary world. In epidemiology, the basic reproduction number, denoted by ℛ_0_, is commonly used to evaluate the transmissibility of an infectious disease like COVID-19. ℛ_0_ is interpreted as the expected number of secondary cases produced by a typical case in a closed population. During the outbreak of an epidemic, ℛ_0_ can be affected by intervention strategies. For example, social measures that decrease the contact rate between individuals would control ℛ_0_. Isolating or treating the infected cases could lower the ℛ_0_ value as well. Another concept the in epidemic theory is the effective reproduction number ℛ_*t*_ which describes the number of people who can be infected by an individual at any specific time *t* in a population. ℛ_*t*_ is time-specific since it accounts for the varying proportions of the population that are immune to the disease over time. There are many recent studies implementing the SIR model (Kermack and McKendrick, 1927) or its modified version to analyze COVID-19 transmissibility in terms of ℛ_0_ or ℛ_*t*_ (see e.g. Chen et al., 2020; Alvarez et al., 2020; Kantner and Koprucki, 2020; Gostic et al., 2020; Cooper et al., 2020). Furthermore, several studies have incorporated the information on social measures to understand the COVID-19 dynamics all over the world. For instance, Dehning et al. (2020) combined the SIR model with Bayesian inference to study the time-varying spreading rate of COVID-19 in Germany. Song et al. (2020) extended the SIR model by considering the quarantine protocols with a focus on understanding the time-course dynamics of COVID-19 in Hubei, China. Giordano et al. (2020) enriched the SIR model with additional compartments to account for under-diagnosis, which could explain the gap between the actual infection dynamics and perception of COVID-19 outbreak in Italy. Because of the heterogeneity in susceptibility and social dynamics, COVID-19 transmissibility differs among locations and changes over time. U.S. local governments have implemented different interventions since mid-March to combat the spread of COVID-19. Therefore, the basic reproduction numbers should spatiotemporally vary.

The basic reproduction number of an epidemic event is changing due to societal and political actions. Effective social measures such as business closures and stay-at-home orders could help lower the transmission rate and thus induce changes in ℛ_0_. By studying the variations in ℛ_0_ over time, we can evaluate the dynamic transmissibility of infectious diseases like COVID-19. For instance, during the outbreak of severe acute respiratory syndrome (SARS) in China around 2003, it was reported an ℛ_0_ ≈ 3.0 for the onset stage of SARS in Hong Kong (Riley et al., 2003; Lloyd-Smith et al., 2003). Later on, it dropped to about 1.1 due to stringent control measures (Chowell et al., 2004). Decreases in ℛ_0_ captured the evolution of SARS transmission dynamics under the approach of efficient diagnosis coupled with patient isolation measures. A recent study in Germany (Dehning et al., 2020) estimated the variations in COVID-19 transmission rates for the four pre-labeled phases partitioned by three time points corresponding to the three major government interventions. Meanwhile, Song et al. (2020) extended the standard SIR model by introducing a transmission rate modifier, which takes different pre-specified decay functions under different macro or micro quarantine measures over time. These studies have enabled public health workers to analyze and evaluate the time-course dynamics of COVID-19 and motivated us to develop a method that can automatically detect the important transitioning time points that occurred during the outbreak of an epidemic, while characterizing the transmission dynamics.

We propose a method named BayesSMILES, which is short for <u>Bayes</u>ian <u>S</u>egmentation <u>M</u>odelIng for <u>L</u>ongitudinal <u>E</u>pidemiological <u>S</u>tudies, to study the dynamics of COVID-19 transmissibility and to evaluate the effectiveness of mitigation interventions. BayesSMILES adopts a Bayesian Poisson segmented regression model to detect multiple change points based on the daily infectious COVID-19 cases. This novel model can 1) capture the epidemiological dynamics under the changing conditions caused by external or internal factors; 2) quantify the uncertainty in both the number and locations of change points; and 3) adjust any relevant explanatory time-varying covariates that may affect the infectious case numbers. In addition, BayesSMILES integrates the change point information to quantify the COVID-19 transmissibility by estimating the basic reproduction numbers in different segments. We demonstrate that our approach can improve the accuracy of the change point detection compared with a widely used change point search method on the simulated data. Applying the proposed BayesSMILES to the U.S. state-level COVID-19 daily report data, we find that the detected change points correlate well with the timing of publicly announced interventions. We also demonstrate that a stochastic SIR model incorporating change point information can provide a better short-term forecast. In all, BayesSMILES enables us to understand the disease dynamics of COVID-19 and provides useful insights for the anticipation and control of current and future pandemics.

The rest of the paper is organized as follows. We review the traditional susceptible-infectious-recovered (SIR) model in Section 2. In Section 3, we describe the framework of BayesSMILES. The Markov chain Monte Carlo (MCMC) algorithm and posterior inference procedures are described in Section 4. We provide a comprehensive simulation study to illustrate the performance of the proposed method against a competing approach in Section 5. In Section 6, we analyze the COVID-19 data for U.S. states using the proposed BayesSMILES. Finally, we conclude with remarks in Section 7 and provide information about implementation in Section 8.

## 2 Review of the SIR Model

The susceptible-infected-removed (SIR) model was developed to simplify the mathematical modeling of human-to-human infectious diseases by Kermack and McKendrick (1927). It is a fundamental compartmental model in epidemiology. At any given time, each individual in a closed population with size *N* is assigned to three distinctive compartments with labels: susceptible (*S*), infectious (*I*), or removed (*R*, being either recovered or deceased). The standard SIR model in continuous time that models the flow of people from *S* to *I* and then from *I* to *R* is described by the following set of nonlinear ordinary differential equations (ODEs):

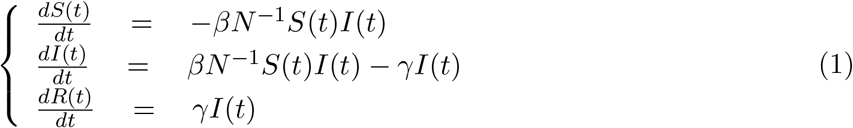

for *t >* 0, subjecting to *S*(*t*) + *I*(*t*) + *R*(*t*) = *N*. Here *β >* 0 is the diseases transmission rate and *γ >* 0 is the removal (recovery or death) rate. Conceptually, susceptible individuals become infectious (*S* → *I*) and then are ultimately removed from the possibility of spreading the disease (*I* →*R*) due to death or recovery with immunity against reinfection.

The rationale behind the first equation in (1) is that the number of new infections during an infinitesimal amount of time, − *dS*(*t*)*/dt*, is equal to the number of susceptible people, *S*(*t*), times the product of the contact rate, *I*(*t*)*/N*, and the disease transmission rate *β*. The third equation in (1) reflects that the infectious individuals leave the infectious population per unit of time, *dI*(*t*)*/dt*, as a rate of *γI*(*t*). The second equation in (1) follows from the first and third ones as a result of *dS*(*t*)*/dt* + *dI*(*t*)*/dt* + *dR*(*t*)*/dt* = 0. Assuming that only a small fraction of the population is infected or removed in the onset phase of an epidemic, we have *S*(*t*)*/N* ≈ 1 and thus the second equation reduces to *dI*(*t*)*/dt* = (*β* − *γ*)*I*(*t*), revealing that the infectious population is growing if and only if *β > γ*. As the expected lifetime of an infected case is given by *γ*^−1^, the ratio *β/γ* is the average number of new infectious cases directly produced by an infected case in a completely susceptible population. Since it is a good indicator of the transmissibility of an infectious disease, the epidemiologists name it the *basic reproduction number* ℛ_0_ = *β/γ* in the context of a standard SIR model, or the *effective reproduction number* ℛ_*t*_ = *β*_*t*_*/γ*_*t*_ in the context of a time-variant SIR model, where *β* and *γ* are replaced by *β*(*t*) and *γ*(*t*) in (1).

The standard SIR model is appealing due to its simplicity. It can be extended in many different ways to better characterize the disease, such as considering vital dynamics, adding more compartments, and allowing more possible transitions between compartments. For instance, the susceptible-exposed-infectious-recovered (SEIR) model includes an additional compartment accounting for the incubation period. The susceptible-infectious-recovered-susceptible (SIRS) model allows recovered individuals to return to a susceptible state. For a comprehensive summary, see Bailey et al. (1975), Becker and Britton (1999), Allen (2008), or Andersson and Britton (2012). It is worth noting that some modified SIR models are currently being used to model the COVID-19 outbreak under under-reporting scenarios (see e.g. Flaxman et al., 2020; Riou et al., 2020; Song et al., 2020).

## 3 The Proposed BayesSMILES Method

### 3.1 Data notations

During a pandemic such as COVID-19, the most accessible and complete data are the daily reported numbers on confirmed cases and deaths. Suppose *N* is the total population size in a given region. Let ***C*** = (*C*_1_, …, *C*_*T*_) and ***D*** = (*D*_1_, …, *D*_*T*_) be the sequences of cumulative confirmed case and death numbers observed at *T* successive equally spaced points in time (e.g. day), where *C*_*t*_ and *D*_*t*_ ∈ ℕ for *t* = 1, …, *T*. For a region for which recovery cases are closely monitored day by day, we use ***E*** = (*E*_1_, …, *E*_*T*_) to denote the sequence of cumulative recovery case numbers. Thus, due to the compositional nature of the basic SIR model, the three trajectories can be constructed as ***S*** = (*S*_1_, …, *S*_*T*_) with *S*_*t*_ = *N − C*_*t*_, ***R*** = (*R*_1_, …, *R*_*T*_) with *R*_*t*_ = *D*_*t*_ + *E*_*t*_, and ***I*** = (*I*_1_, …, *I*_*T*_) with *I*_*t*_ = *N* − *S*_*t*_ − *R*_*t*_ = *C*_*t*_ − *D*_*t*_ − *E*_*t*_. For a region for which recovery cases do not exist or are under-reported, we consider both ***R*** and ***I*** as missing data and reconstruct these two sequences according to the last equation of (1) with a pre-specified constant removal rate *γ*. Specifically we set *I*_1_ = *C*_1_ and *R*_1_ = 0, and generate *R*_*t*_ = *R*_*t*−1_ + *γI*_*t*−1_ and *I*_*t*_ = *I*_*t*−1_ + (*C*_*t*_ − *C*_*t*−1_) − (*R*_*t*_ − *R*_*t*−1_) from *t* = 2 to *T* sequentially, where ⌈·⌉ : ℝ^+^ →N denotes the ceiling function. For the choice of *γ*, we suggest estimating its value from publicly available reports in some region where confirmed, death, and recovery cases are all well-documented, or from prior epidemic studies due to the same under-reporting issue in actual data. Lastly, given a vector ***Y*** = (*Y*_1_, …, *Y*_*T*_) and some initial value *Y*_0_ (for example, ***Y*** can be ***C, D, E, S, I*** or ***R***), we use 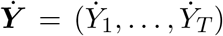 to denote the lag one difference of ***Y***, where 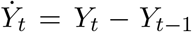 for *t* = 1, …, *T*; that is, 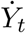 is the difference between two adjacent observations. Table 1 and 2 summarize the data notations as well as the key notations of models introduced in Section 3.3 and 3.4, respectively.

**Table 1:** Key notations of the Poisson segmented regression model described in Section 3.3

|  | Notation | Support | Definition |
| --- | --- | --- | --- |
| Data | $N$ | $N \in \mathbb{N}$ | The total population size |
| | $\mathbf{C} = [C_t]_{T \times 1}$ | $C_t \in \mathbb{N}, C_t \geq C_{t-1}$ | The cumulative confirmed case numbers |
| | $\mathbf{D} = [D_t]_{T \times 1}$ | $D_t \in \mathbb{N}, D_t \geq D_{t-1}$ | The cumulative death numbers |
| | $\mathbf{E} = [E_t]_{T \times 1}$ | $E_t \in \mathbb{N}, E_t \geq E_{t-1}$ | The cumulative recovery case numbers |
| | $\mathbf{S} = [S_t]_{T \times 1}$ | $S_t \in \mathbb{N}$ | The actively susceptible people, $S_t = N - C_t$ |
| | $\mathbf{I} = [I_t]_{T \times 1}$ | $I_t \in \mathbb{N}$ | The actively infectious case numbers |
| | $\mathbf{R} = [R_t]_{T \times 1}$ | $R_t \in \mathbb{N}, R_t \geq R_{t-1}$ | The cumulative removed case numbers |
| | $\mathbf{X} = [x_{t,j}]_{T \times p}$ | $x_{t,j} \in \mathbb{R}, j > 2$ | The design matrix, where the first column is an all-one vector for the intercept, the second column is $(1, \dots, T)^\top$ for the time effect, and all other columns make up the covariate matrix |
| | $\gamma$ | $\gamma \in \mathbb{R}^+$ | The pre-specified constant removal rate |
| | $\boldsymbol{\sigma}^2 = [\sigma_k^2]_{K \times 1}$ | $\sigma_k^2 \in \mathbb{R}^+$ | The pre-specified variances of regression errors |
| Parameters | $\boldsymbol{\beta} = [\beta_t]_{T \times 1}$ | $\beta_t \in \mathbb{R}^+$ | The time-varying disease transmission rates |
| | $\boldsymbol{\alpha} = [\alpha_t]_{T \times 1}$ | $\alpha_t \in \mathbb{R}^+$ | The adjusted time-varying transmission rates |
| | $\tilde{\boldsymbol{\alpha}} = [\tilde{\alpha}_t]_{T \times 1}$ | $\tilde{\alpha}_t \in \mathbb{R}$ | The adjusted time-varying transmission rates on the logarithmic scale, $\tilde{\alpha}_t = \log \alpha_t$ |
| | $\boldsymbol{\zeta} = [\zeta_t]_{T \times 1}$ | $\zeta_t \in \{0, 1\}$ | The change point indicator |
| | $\mathbf{z} = [z_t]_{T \times 1}$ | $z_t \in \{1, \dots, K\}$ | The segmentation indicator |
| | $\mathbf{c} = [c_k]_{K \times 1}$ | $c_k \in \{1, \dots, T\}$ | The locations of change points |
| | $\mathbf{n} = [n_k]_{K \times 1}$ | $n_k \in \mathbb{N}$ | The number of time points in each segment |
| | $\mathbf{B} = [b_{j,k}]_{p \times K}$ | $b_{j,k} \in \mathbb{R}$ | The regression coefficients for each segment |
| | $\boldsymbol{\epsilon} = [\epsilon_t]_{T \times 1}$ | $\epsilon_t \in \mathbb{R}$ | The regression errors (i.e. the process error) |
| | $\mathbf{H} = \text{Diag}(h_0, \dots, h_{p-1})$ | $h_j \in \mathbb{R}^+$ | The diagonal variance-covariance matrix that defines the prior on each column in $\mathbf{B}$, i.e. $\mathbf{b}_k$ |
| | $\omega$ | $\omega \in (0, 1)$ | The probability of being a change point *a priori* |
| | $a_\omega, b_\omega$ | $a_\omega, b_\omega \in \mathbb{R}^+$ | The hyperparameters for $\omega$ |
| Others | $\top$ | | The matrix transpose operator |
| | $\lceil \cdot \rceil$ | | The ceiling function |
| | $\delta(\cdot)$ | | The indicator function |
| | $\mathcal{I}_n = \text{Diag}(1, \dots, 1)$ | | The $n$-by-$n$ identity matrix |
| | $\mathbf{0}_n = [0]_{n \times 1}$ | | The $n$-dimension all-zero column vector |

**Table 2:**
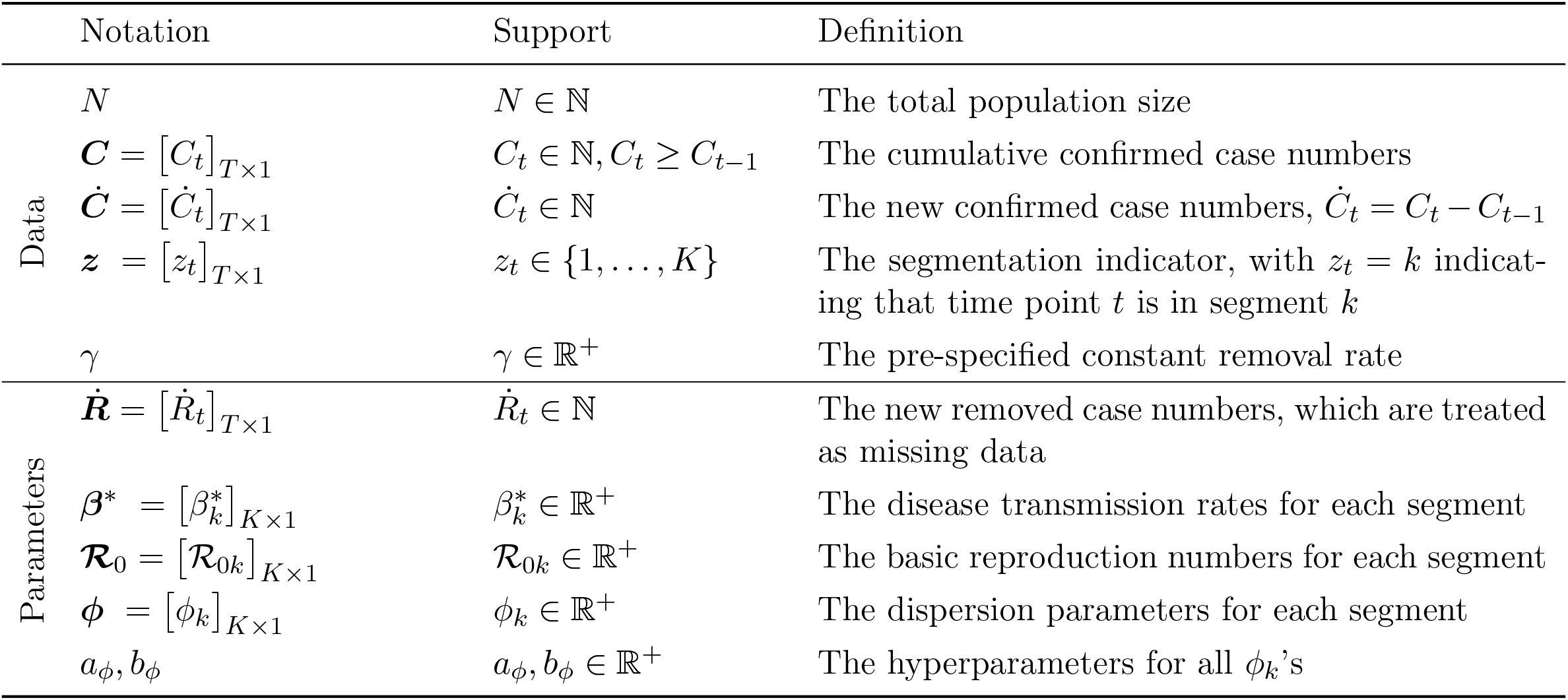
Key notations of the stochastic SIR model described in Section 3.4

### 3.2 Modeling epidemic dynamics via a modified stochastic SIR model

An SIR model has three trajectories, one for each compartment. The compositional nature of the three trajectories, i.e. *S*(*t*) + *I*(*t*) + *R*(*t*) = *N*, implies that we need only two of them to solve the ODEs as shown in (1). As mentioned previously, assuming *S*(*t*) ≈ *N* for all *t* results in *dI*(*t*)*/dt* = (*β* − *γ*)*I*(*t*) and further leads to an exact solution: *I*(*t*) = *I*(0) exp {(*β* − *γ*)}*t*. For modeling daily reported actively infectious data ***I***, we utilize its discrete-time version,

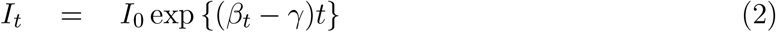

with a time-varying rate *β*_*t*_ to account for the transmissibility changes of the disease. For simplicity’s sake, we assume a constant removal rate *γ*. Based on (2), we introduce a probabilistic model, which approximately mimics the dynamics of the deterministic standard SIR model as shown in (1) during the onset phase of a pandemic. Specifically, we suppose the infectious population size at time *t* is sampled from a Poisson model,

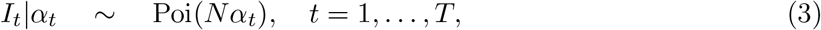

where *α*_*t*_ = *I*_0_ exp {(*β* − *γ*)*t*}/*N* is a redefined time-varying transmissibility parameter that depends on the initial infectious population size *I*_0_, the disease transmission rate *β*_*t*_, the removal rate *γ*, and any latent factors (e.g. public health intervention, social behavior, virus mutation, healthcare quality, etc.) that may affect the disease transmissibility. This model automatically accounts for measurement errors and uncertainties associated with the counts. Note that (3) can be generalized to a negative binomial (NB) model, i.e. *I*_*t*_|*α*_*t*_ ∼ NB(*Nα*_*t*_, *ϕ*_*I*_) if needed, where *ϕ*_*I*_ is a dispersion parameter aiming to account for over-dispersion that might be observed in ***I***. Here we use NB(*µ, ϕ*), *µ, ϕ >* 0 to denote an NB distribution with expectation *µ* and variance *µ* + *µ*^2^*/ϕ*.

### 3.3 Detecting change points via a Poisson segmented regression model

Our change point detection builds upon the above modified stochastic time-variant SIR model (3) with stationary transmissibility *α*_*t*_ in a certain segment. Particularly, we assume that *β*_*t*_ only changes at certain time points. Identifying those change points is of significant importance, as it not only enables us to characterize the temporal dynamics of the pandemic but also helps policymakers evaluate the effectiveness of the past and ongoing mitigation and intervention strategies.

In this paper, the change points are defined as those discrete time points that significantly alter the disease transmission rate *β*_*t*_ between two adjacent segments, given a constant removal rate *γ* across all time points. We introduce a latent binary vector ***ζ*** = (*ζ*_1_, …, *ζ*_*T*_), *ζ*_*t*_ ∈ {0, 1}, with *ζ*_*t*_ = 1 if time point *t* is a change point and *ζ*_*j*_ = 0 otherwise. We set *ζ*_1_ = 1 by default, interpreting the first time point as the “zeroth change point.” Those points with *ζ*_*j*_ = 0 can be partitioned into segments bounded by two adjacent change points. Thus, we use another vector ***z*** = (*z*_1_, …, *z*_*T*_), *z*_*t*_ ∈ {1, …, *K*} to reparameterize ***ζ***, where we define 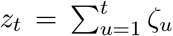. Thus, *z*_*t*_ = *k* indicates that time point *t* is in segment *k*, that is, between the (*k* − 1) and *k*-th change points. The total number of change points excluding the first time point is *K* − 1. Note that ***ζ*** is the lag one difference of ***z***, i.e. *ζ*_*t*_ = *z*_*t*_ − *z*_*t*−1_ with *ζ*_1_ = 1. Figure 1 shows the representations of ***ζ*** and ***z*** for a simulated time-series dataset (*T* = 10) with two change points.

**Figure 1:**
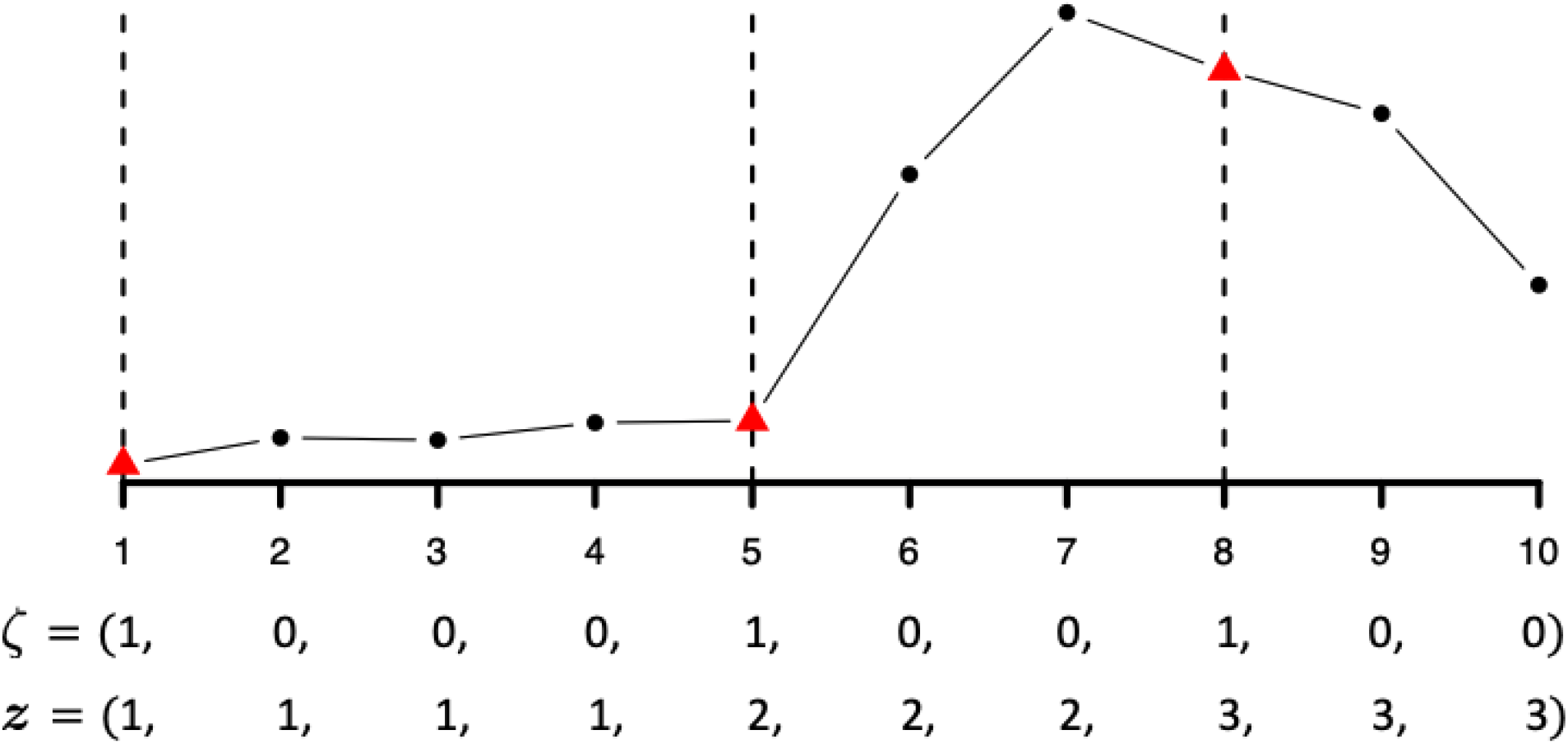
An example of time-series data (*T* = 10) with two change points (*K* = 3) and its associated parameterizations in terms of ***ζ*** and ***z***, respectively. Red triangles are change points, while black circles are not. Note that the first time point is treated as the “zeroth change point.”

To infer ***ζ*** or ***z*** given the sequence of infectious population size ***I***, we adopt a Poisson segmented regression framework, which can be written as,

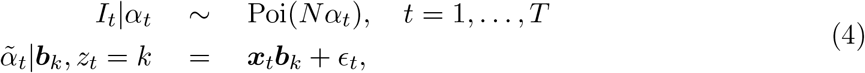

where 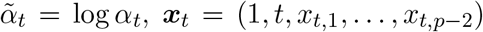 is a *p*-dimensional row vector of covariates that includes a scalar of one for the intercept, time *t*, and *p* − 2 explanatory variables observed at time *t* if applicable. Those explanatory variables could contain the number of tests, weather information, mobility report, or other necessary and accessible time-varying measures important to adjust for during a longitudinal epidemiological study. The vector ***b***_*k*_ = (*b*_1,*k*_, …, *b*_*p,k*_)^*T*^ is a *p*-dimensional column vector of segment-specified coefficients that includes an intercept representing the proportion of infectious people at logarithmic scale, i.e. *b*_1,*k*_ = log(*I*_0_*/N*), in segment *k*, and a slope accounting for the time-varying disease transmission rate, i.e. *b*_2,*k*_ = *β*_*t*_−*γ*.

Let ***X*** denote the design matrix, which combines all ***x***_*t*_’s as rows and ***B*** denote the corresponding coefficient matrix, which combines all ***b***_*k*_’s as columns. For simplicity’s sake, we assume the process error _1_, …, _*T*_ are independent and identically Gaussian distributed with zero mean and segment-specified variance, i.e. 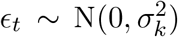. To ensure the identifiability of all model parameters, we try a set of considerably small values for 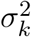’s and employ a robust cross validation method called Pareto-smoothed importance sampling leave-one-out (PSIS-LOO) cross validation to determine the best choice (Vehtari et al., 2017).

Let ***α***_*k*_ be the sequence of all *α*_*t*_’s in segment *k*, i.e. 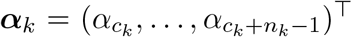, where we denote *c*_*k*_ = min{*t* : *z*_*t*_ = *k*} as the location of the (*k*−1)-th change point and 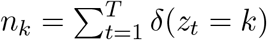 as the number of time points in segment *k* with *δ*(·) being the indicator function. We can rewrite the second equation in (4) as 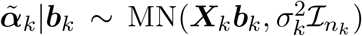, where 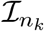 is an *n*_*k*_-by-*n*_*k*_ identity matrix and ***X***_*k*_ can be explicitly written as

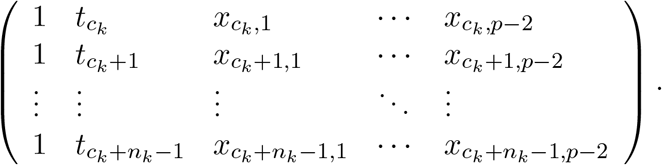

We assume a zero-mean multivariate normal distribution for ***b***_*k*_, that is, ***b***_*k*_ ∼ MN(**0**_*p*_, ***H***), where **0**_*p*_ is an *p*-by-1 all-zero column vector and ***H*** = Diag(*h*_0_, …, *h*_*p*−1_) is set to be a diagonal variance-covariance matrix. For a weakly informative setting, we recommend choosing a large value for each *h*_*j*_. Through this prior specification, we are able to marginalize out the nuisance parameter ***b***_*k*_ and obtain 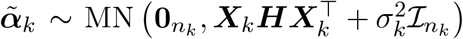. Consequently, we can write the collapsed model of (4) as

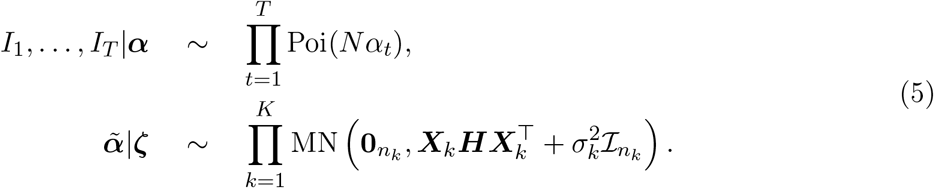

To complete the model specification, we impose an independent Bernoulli prior on *ζ* as 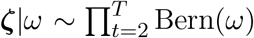 Bern(*ω*), where *ω* is interpreted as the probability of a time point being a change point *a priori*. We further relax this assumption by allowing *ω* ∼ Be(*a*_*ω*_, *b*_*ω*_) to achieve a beta-Bernoulli prior. In practice, we suggest a constraint of *a*_*ω*_ + *b*_*ω*_ = 2 for a vague hyperprior of *ω* (Tadesse et al., 2005). In addition to that, we make the prior probability of ***ζ*** equal to zero if two adjacent time points are jointly selected as change points. In other words, a segment should consist of at least two time points.

### 3.4 Estimating basic reproduction numbers via a stochastic SIR model

Given the multiple change points ***ζ***, we estimate the basic reproduction number ℛ_0_ = *β/γ* for each segment via a stochastic version of the standard SIR model as shown in (1), which only needs the cumulative confirmed case numbers ***C***. This is because recovery data exist in only a few states in the U.S., which makes both model inference and predictions infeasible. This model considers both of the removed and actively infectious cases as missing data and mimics their relationship as in some compartmental models in epidemiology. Specifically, we assume the number of new removed cases at time *t*, i.e. 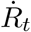, is sampled from a Poisson distribution with mean *γI*_*t*−1_, that is, 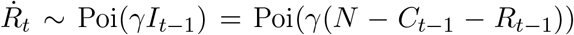, where *γ* should be pre-specified. Based on this simplification, we rewrite the discrete version of the first equation in (1) as,

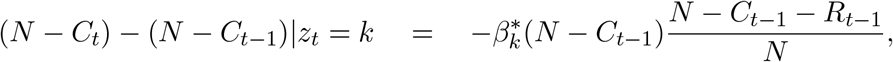

resulting in

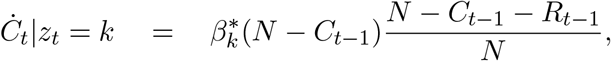

where 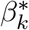 is the common disease transmission rate for the all the time points in segment *k*.

We further assume the new case number observed at time *t*, i.e. *Ċ*_*t*_, is sampled from an NB model,

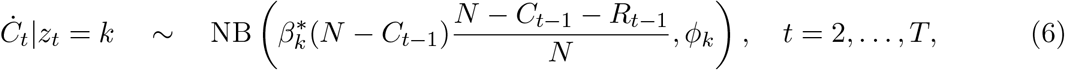

as it automatically accounts for measurement errors and uncertainties associated with the counts. Following most epidemiological models, we assume this stochastic process is a Markov process, where the present state (at time *t*) depends only upon its previous state (at time *t* − 1). The setting above builds upon the standard SIR model. It is worth noting that the oversimplified assumptions of the proposed stochastic SIR model, as well as the bias in data reporting, might undermine the reliability of the estimates on disease transmission rates 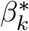 s and their succeeding basic reproduction numbers ℛ_0*k*_’s. However, they can still be used as a proxy to indicate the transmissibility dynamic of an infectious disease. We could consider additional compartments as seen in the susceptible-infectious (SIS) model, the susceptible-infectious-recovered-deceased (SIRD) model, the susceptible-exposed-infectious-removed (SEIR) model, and the susceptible-exposed-infectious-susceptible (SEIS) model (see a comprehensive summary in Bailey et al. (1975)). The effects from the additional compartments could be incorporated by reparameterizing the mean function in the NB distribution, as shown in Equation (6), which is left as future work. For the prior distribution of the segment-specific dispersion parameter *ϕ*_*k*_, we choose a gamma distribution, *ϕ*_*k*_ ∼ Ga(*a*_*ϕ*_, *b*_*ϕ*_) for *k* = 1, …, *K*. We recommend small values, such as *a*_*ϕ*_ = *b*_*ϕ*_ = 0.001, for a weakly informative setting. This model, on average, mimics the epidemic dynamics and is more flexible than those deterministic epidemiological models. For each segment *k*, we assume 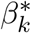 comes from a gamma distribution with hyperparameters that makes both mean and variance of the transformed variable 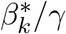 equal to 1. Table 2 summarizes the notations for the model parameters described above.

## 4. Model Fitting

In this section, we describe the Markov chain Monte Carlo (MCMC) algorithms for posterior inference of the proposed BayesSMILES method, including the inferential strategy for identifying change points and estimating the basic reproduction numbers, respectively.

### 4.1 MCMC algorithms for detecting change points

Our primary interest lies in identifying the change point locations via the vector ***ζ*** based on the actively infectious cases ***I***. According to Section 3.3, the full data likelihood and the priors of the change point detection model are written as,

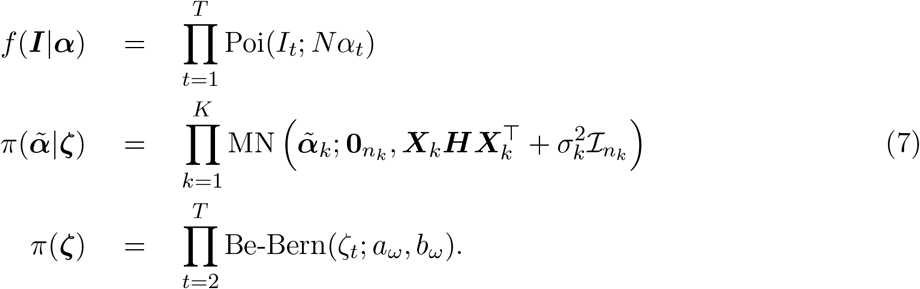

Thus, the full posterior distribution is 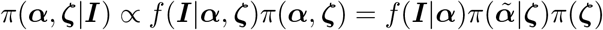. Since there are no closed form expressions for the two conditionals *π*(***α*** | ***ζ, I***) and *π*(***ζ*** | ***α, I***), we use Metropolis-Hastings (MH) algorithms to sample from the two distributions.

#### Update the change point indicator *ζ*

We update the binary latent vector ***ζ*** via an *add-delete-swap* algorithm. We randomly select an entry in ***ζ***, say *ζ*_*t*_, and change its value to 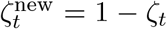 to form a new *ζ*^new^. This is an *add* step if 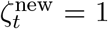 and a *delete* step otherwise. The *swap* step is performed every ten iterations, where we randomly select a change point, say *ζ*_*t*_ = 1, and swap the values between the *t* and (*t* ± 1)-th entries in ***ζ*** to form a new ***ζ***^new^. We accept the proposed ***ζ***^new^ with the probability min(1, *m*_MH_), where the acceptance ratio is

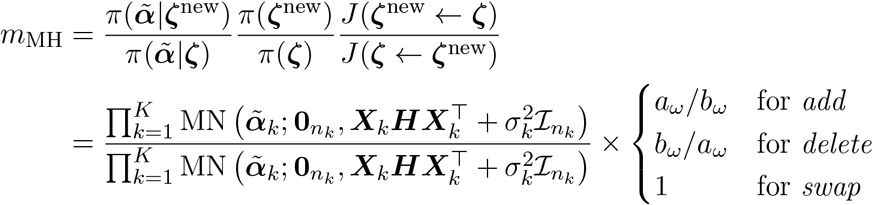

Here we use *J* (· ← ·) to denote the proposal probability distribution for the selected move. Note that the last proposal density ratio equals one. This step simultaneously updates the segmentation vector ***z***, as it can be constructed from ***ζ***.

#### Update the adjusted time-varying transmission rates *α*

For each segment partitioned by ***ζ***, we update *α*_*t*_ within the same segment, say segment *k*, sequentially by using a random walk Metropolis-Hastings (RWMH) algorithm. We first propose a new 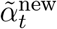 from 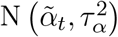. Let 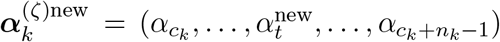. Then we accept the proposed value 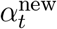 with probability min(1, *m*_MH_), where the acceptance ratio is

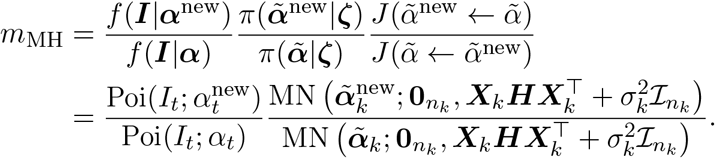

Note that the proposal density ratio cancels out for this RWMH update. The computation of the multivariate normal (MN) probability density involves matrix inversion, which can be time-consuming, particularly when *n*_*k*_ is large. To significantly improve the computational efficiency, we follow Zhou and Guan (2018) to approximate the exact inversion under an appropriate choice of ***H*** that satisfies the asymptotic condition. As mentioned previously, ***H*** is a *p*-by-*p* diagonal matrix, where the first entry *h*_0_ corresponds to the variance of the normal prior on *b*_1,*k*_. Under the asymptotic condition of *h*_0_ ≥ *h*_*j*_, ∀ *j* ≠ 0, the inversion of an *n*_*k*_-by-*n*_*k*_ matrix is reduced to an inversion of a *p*-by-*p* matrix (See more details in Appendix A1). In practice, we set *h*_0_ = 10, 000 and *h*_1_ = … = *h*_*p*−1_ = 10 to ensure this asymptotic condition. The full details of the approximation method are available in the Appendix A1.

### 4.2 MCMC algorithms for estimating basic reproduction numbers

Once the change points are determined, we aim to estimate the basic reproduction numbers ℛ_0_’s across different segments and quantify their uncertainties based on the cumulative confirmed cases ***C*** only. According to Section 3.4, the full data likelihood and the priors of the stochastic SIR model are written as,

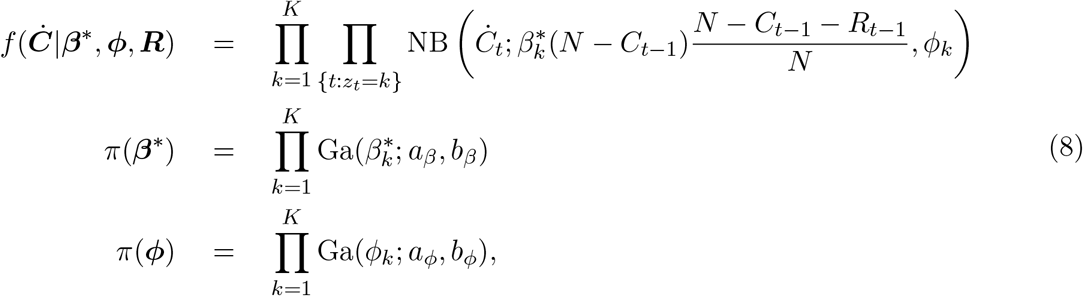

where 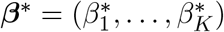 and ***ϕ*** = (*ϕ*_1_, …, *ϕ*_*K*_), i.e. the collections of transmission and dispersion rates of all segments. For the hyperparameters, we set *a*_*β*_ = 1 and *b*_*β*_ = 1*/γ* so that both of the expectation and variance of the basic reproduction number 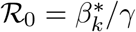 are equal to one. With a pre-defined removal rate *γ*, we propose the following updates in each MCMC iterations.

#### Generate *R* based on *C*

We assume *I*_1_ = *C*_1_ and *R*_1_ = 0, i.e. all the confirmed cases are capable of passing the disease to all susceptible individuals in a closed population at time point *t* = 1. Then we sample 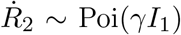, where *γ* is a pre-specified tuning parameter and 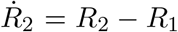 is the new removed case number at time point *t* = 2. Due to the compositional nature of the SIR model, we can compute 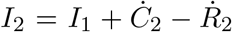, where *Ċ*_2_ = *C*_2_ − *C*_1_ is the new confirmed cases at time point *t* = 2. Next, we repeat this process of sampling 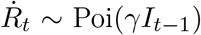 and computing 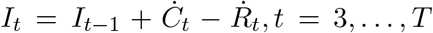, to generate the sequence ***R*** used in every iteration.

#### Update the dispersion parameters *ϕ*

For each segment, we update *ϕ*_*k*_ by using an RWMH algorithm. We first propose a new 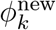, of which the logarithmic value is generated from 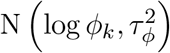. Let 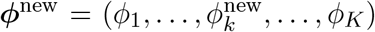, where only the *k*-th entry is replaced. Then we accept the proposed value 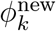 with probability min(1, *m*_MH_), where the acceptance ratio is

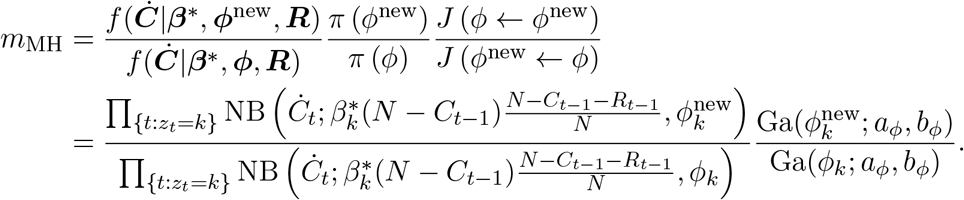

Note that the proposal density ratio cancels out for this RWMH update.

#### Update the disease transmission rates *β*^*^

For each segment, we update 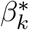by using an RWMH algorithm. We first propose a new 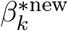, of which the logarithmic value is generated from 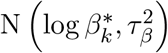. Let 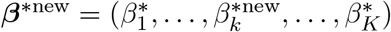, where only the *k*-th entry is replaced. Then we accept the proposed value 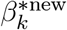 with probability min(1, *m*_MH_), where the acceptance ratio is

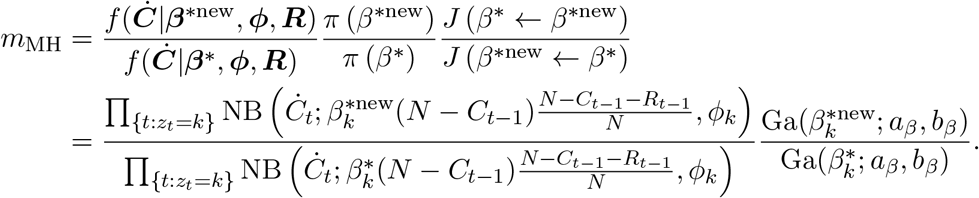

Note that the proposal density ratio cancels out for this RWMH update.

### 4.3 Posterior inference

We explore posterior inference for the parameters of interest by postprocessing the MCMC samples after burn-in iterations. We start by obtaining a point estimate of the change point indicator ***ζ*** by analyzing its MCMC samples {***ζ***^(*u*)^, …, ***ζ***^(*U*)^}, where *u* indexes the MCMC iteration after burn-in. One way is to choose the ***ζ*** corresponding to the *maximum-a-posteriori* (MAP),

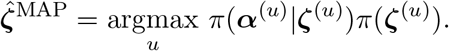

The corresponding 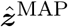 can be obtained by taking the cumulative sum of 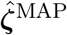. An alternative estimate relies on the computation of posterior pairwise probability matrix (PPM), where the probability that time points *t* and *t*′ are assigned into the same segment is estimated by 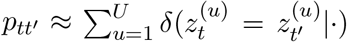. This estimate utilizes the information from all MCMC samples and is thus more robust. After obtaining this *T* -by-*T* co-clustering matrix denoted by ***P*** = [*p*_*tt*_′]_*T ×T*_, a point estimate of ***z*** can be approximated by minimizing the sum of squared deviations of its association matrix from the PPM, that is,

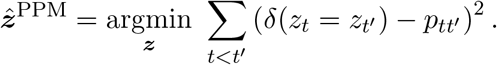

The corresponding 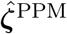 can be obtained by taking the difference between consecutive entries in 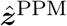 and setting the first entry to one. To construct a “credible interval” for a change point, we utilize its local dependency structure from all MCMC samples of ***ζ*** that belong to its neighbors. Due to the nature of the MCMC algorithm described in Section 4.1, if a time point *t* is selected as a change point, i.e. *ζ*_*t*_ = 1, then its nearby time points must not be a change point. Thus, the correlation between the MCMC sample vectors 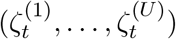 and 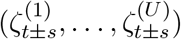 tends to be negative when *s* is small. We define the credible interval of a change point as the two ends of all its nearby consecutive time points, for which the MCMC samples of ***ζ*** are significantly negatively correlated with that of the change point. This could be done via a one-sided Pearson correlation test with a pre-specified significant level, e.g. 0.05. Although quantifying uncertainties of change points is not rigorous, it performs very well in the simulation study and yields reasonable results in the real data analysis.

Once the change points are determined, an approximate Bayesian estimator of the disease transmission rate 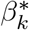 for each segment *k* can be simply obtained by averaging over all of its MCMC samples, 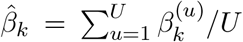. In addition, a quantile estimation or credible interval can be obtained. Lastly, we summarize the basic reproduction number in each segment *k* as 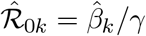.

### 4.4 Prediction

Conditional on the change point locations, we can predict the cumulative or new confirmed cases at any future time *T*_*f*_ by Monte Carlo simulation based on the information in the last segment *K* only. Specifically, from time *T* + 1 to *T*_*f*_, we sequentially generate

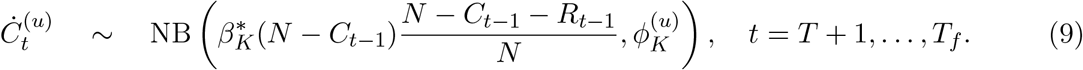

Then, both short and long-term forecasts can be made by summarizing the (*T*_*f*_ − *T*)-by-*U* matrix of MCMC samples. For instance, the predictive number of cumulative and new confirmed cases at time *T* + 1, on average, are 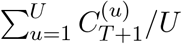 and 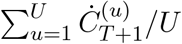, respectively.

## 5 Simulation

We used simulated data to evaluate the performance of our BayesSMILES method in terms of both change point detection and basic reproduction number estimation. It is shown that the proposed Bayesian framework outperforms an alternative change point detection method.

### 5.1 The generative model

The three trajectories ***S, I***, and ***R*** with length *T* = 120 were generated in the following way. We first divided the *T* = 120 time points into *K* = 4 segments with the same length; that is, the true change points were *t* = 31, *t* = 61, and *t* = 91. To mimic the disease transmissibility dynamics across different segments, we chose segment-varying disease transmission rates 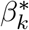 while fixing the removal rate *γ* = 0.03. Let ℛ_0_ be a *K*-vector where each entry gives the reproduction number of one segment, which can be computed by 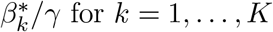 for *k* = 1, …, *K*. We considered four scenarios of the set 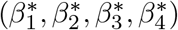, corresponding to 1) ℛ_0_ = (3.0, 1.2, 2.0, 0.8); 2) ℛ_0_ = (3.0, 2.3, 1.5, 0.8); 3) ℛ_0_ = (3.0, 1.8, 0.8, 1.6); 4) ℛ_0_ = (3.0, 2.0, 1.1, 0.5). Then based on the stochastic version of the standard SIR model, we sampled *S*_*t*_ and *R*_*t*_ from negative binomial (NB) distributions, and obtained *I*_*t*_, sequentially from *t* = 1 to *T* through

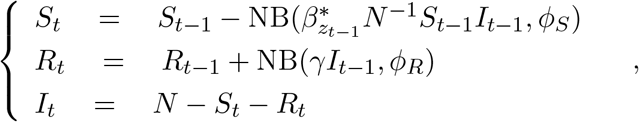

where *N* = 1, 000, 000, the initial *I*_0_ = 100 and *R*_0_ = 0, and the dispersion parameters *ϕ*_*S*_ = *ϕ*_*R*_ = 10. Note that the generative scheme was with an NB error structure, which was different from our model assumption based on a Poisson error structure. We repeated the above steps to generate 50 independent datasets for each setting of ℛ_0_. Figure 2 displays the temporal patterns of the simulated infectious counts ***I*** for the four scenarios.

**Figure 2:**
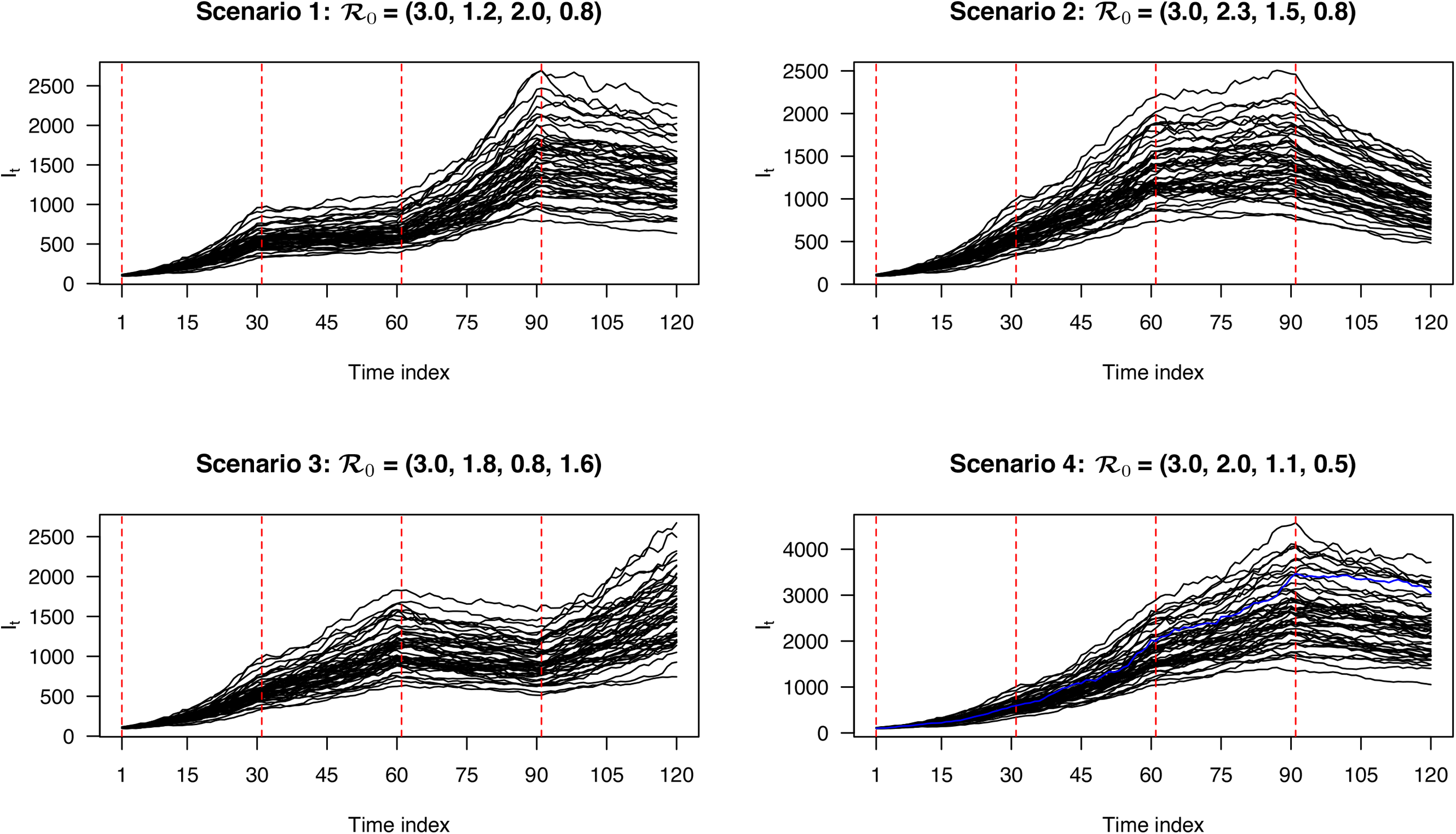
Simulation study: The simulated actively infectious data ***I*** under the four scenarios. Each curve represents a replicated sequence of ***I*** under a scenario. The red dashed lines mark the true change point locations. The blue curve under Scenario 4 was randomly chosen for evaluating the model fitting, of which results are shown in Figure 3.

### 5.2 Evaluation criteria

To evaluate the change point detection, we may rely on either the binary change point indicator vector ***ζ*** or the time point allocation vector ***z***. For the choice of ***ζ***, a change point is considered to be correctly identified if its location is within a local window of the true position (Killick and Eckley, 2014). The selection of the window size is *ad hoc* and may bias the evaluation. In addition to that, since change points and non-change points are usually of very different sizes, most of the binary classification metrics are not suitable for model comparison here. Thus, we chose those metrics that quantify the agreement between the true and estimated allocation vectors, i.e. ***z*** and 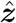. The two classic performance metrics for the analysis of clustering results are the adjusted Rand index (ARI) and mutual information (MI), proposed by Hubert and Arabie (1985) and Steuer et al. (2002), respectively. ARI is the corrected-for-chance version of the Rand index (Rand, 1971), as a similarity measure between two sample allocation vectors. Let 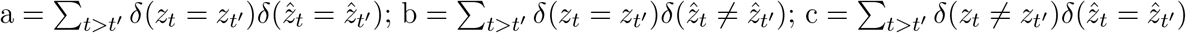 and 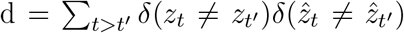 be the number of pairs of time points that are a) in the same segment in both of the true and estimated partitions; b) in different segments in the true partition but in the same segment of the estimated one; c) in the same segment of the true partition but in different segments in the estimated one; and d) in different segments in both of the true and estimated partitions. Then, the ARI can be computed as

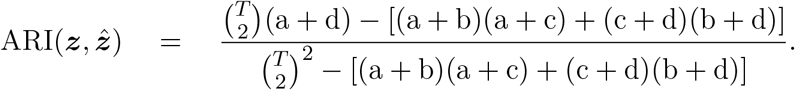

The ARI usually yields values between 0 and 1, although it can yield negative values (Santos and Embrechts, 2009). The larger the index, the more similarities between ***z*** and 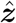, and thus the more accurately the method detects the actual times at which change points occurred. An alternative metric choice is MI, which measures the information about one variable that is shared by the other (Steuer et al., 2002). Let 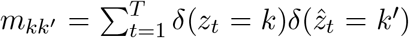 be the number of time points shared between the *k*-th segment in the true ***z*** and the *k*^*i*^-th segment in the estimated one 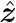. Then, MI can be computed as

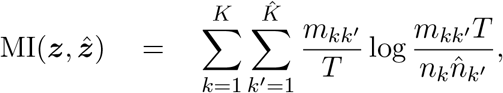

where 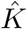 is the number of segments and 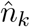s are the segment lengths for segment 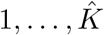 in 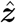. It yields non-negative values. The larger the MI, the more accurate the partition result.

To quantify how well a method estimates the dynamic transmissibility across different segments, we used the root mean square error (RMSE) that measures the deviation between the true and estimated values of ℛ_0_ over all *T* time points:

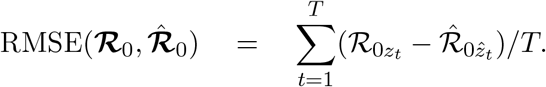

A smaller value of RMSE indicates a more accurate estimation of ℛ_0_’s.

### 5.3 Results

As for the MCMC setting of change point detection, we set 40, 000 MCMC iterations and discarded the first half as burn-in. We adopted the weakly informative setting by setting *a*_*ω*_ = 0.1 and *b*_*ω*_ = 1.9 in the Beta-Bernoulli prior for the change point indicator vector ***ζ***. We set ***H*** = Diag(*h*_0_, *h*_1_) with *h*_0_ = 10, 000 and *h*_1_ = 10 as the covariance matrix in the prior distribution of ***b***_*k*_’s. Finally, we let 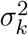 take ten equally spaced values ranging from 0.0001 to 0.01 at the logarithmic scale (base 10) in the PSIS-LOO cross validation. In fitting the stochastic SIR model, we set 100, 000 MCMC iterations with the first half as burn-in. As suggested in Waqas et al. (2020), the value of removal rate *γ* could be estimated by 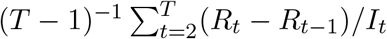 for each simulated dataset. Then, we set *a*_*β*_ = 1 and *b*_*β*_ = 1*/γ* so that both the prior expectation and prior variance of the basic reproduction number 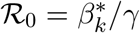 are equal to 1.

We first checked the performance of BayesSMILES on a single simulated dataset, which was randomly selected from the 50 replicates in Scenario 4 (marked as the blue line in Figure 2). Note that we did the same for the remaining three scenarios, and the related results are summarized in Appendix A2. Figure 3(a) demonstrates the change point detection result based on the Poisson segmented regression model. The red dashed and the blue solid lines represent the true and the estimated change point locations, respectively, while the gray ribbons represent the 95% credible intervals for those identified change points. As we can see, BayesSMILES successfully detected the three true change points in general, as each of the 95% credible intervals covered the truth. The resulted values of ARI and MI were 0.93 and 1.28, respectively. Later on, the stochastic SIR model introduced in Section 3.4 was then fitted to quantify the disease transmissibility in each segment bounded by the identified change points. Figure 3(b) shows the posterior distributions of 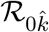’s for 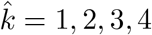 from their MCMC samples. The red dashed and blue solid lines pinpoint the true and posterior mean of 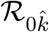’s, while the two black solid lines mark the boundary of their 95% credible intervals. Clearly, those true values were within their corresponding 95% credible intervals. The final RMSE for ℛ_0_ estimation was 0.38 for this single simulated dataset.

**Figure 3:**
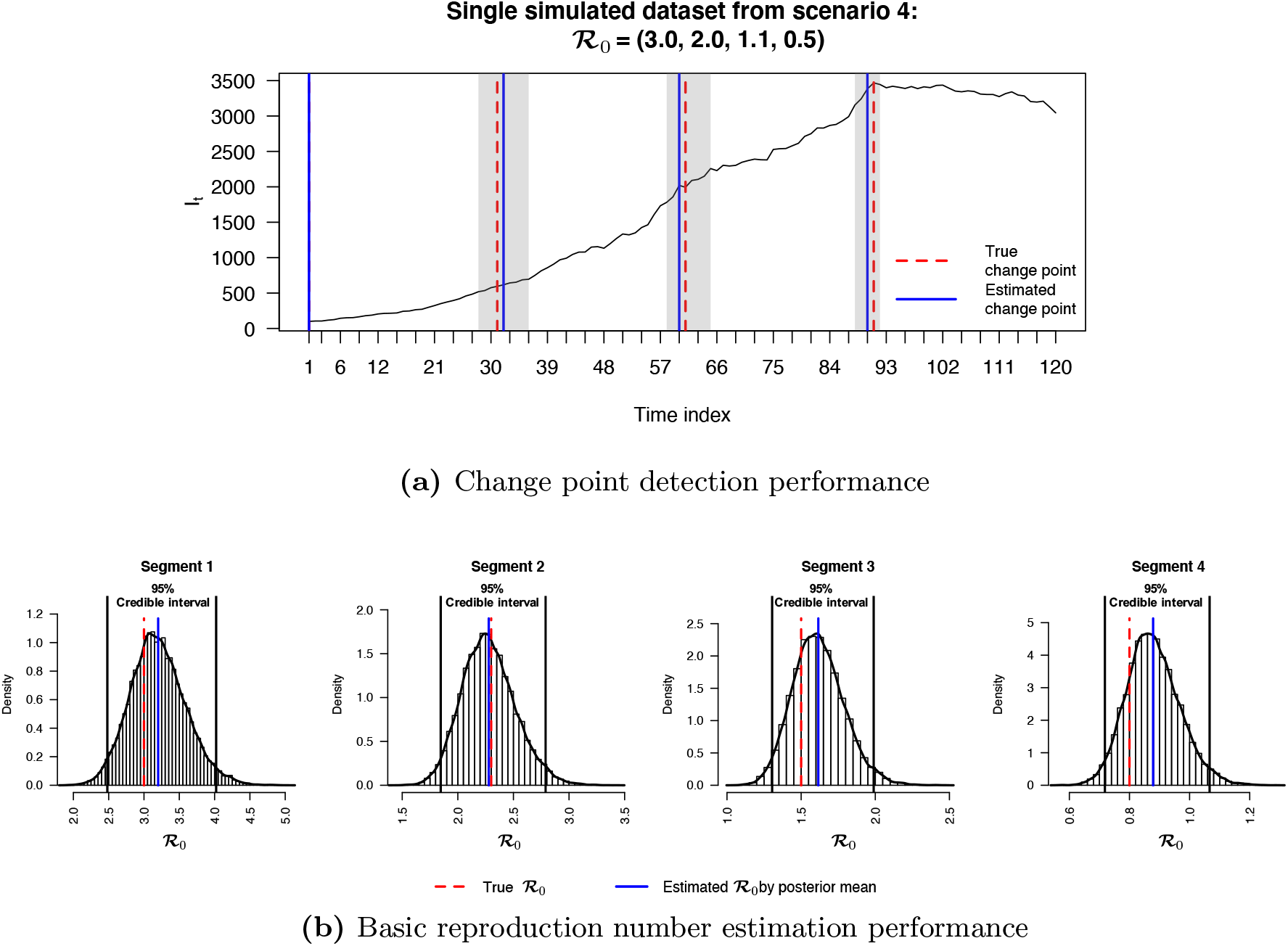
Simulation study: The model fitting results based on a randomly selected simulated dataset (see the blue curve under Scenario 4 in Figure 2). (a) The locations of change points (blue solid lines) estimated from the posterior pairwise probability matrix (PPM) and their credible intervals (gray ribbons). The red dashed lines mark the true change point locations; (b) The posterior distributions of ℛ_0*k*_’s for *k* = 1, 2, 3, 4 estimated from the segmented time-series data, given the three identified change points as shown in (a). The red dashed and blue solid lines are the true and estimated values of ℛ_0*k*_’s, respectively. The two black solid lines are the lower and upper bounds of the 95% credible intervals.

To the best of our knowledge„ there is no method like BayesSMILES that can detect latent change points while characterizing the transmission dynamics through an SIR model. Thus, in setting up a comparison study, we therefore considered a two-stage approach that first identifies multiple change points of time-series data based on a likelihood based framework, and then estimates the basic reproduction numbers between each pair of nearby change points, following the stochastic SIR model introduced in Section 3.4. The alternative change point model assumes time points within one segment follow a normal distribution with distinct mean and/or variance from its nearby segments (Hinkley, 1970; Jen and Gupta, 1987), and it uses the likelihood ratio test (LRT) to detect multiple change points. An algorithm named binary segmentation (Edwards and Cavalli-Sforza, 1965; Sen and Srivastava, 1975) is commonly used to compute the test statistics for the LRT with high efficiency (Killick et al., 2012). In our case, to detect change points using this alternative approach named the likelihood ratio test with binary segmentation (LRT-BinSeg), we input the logarithmic scale of ***I*** into the function cpt.meanvar in the related `R` package `changepoint` (Killick and Eckley, 2014) for each of the simulated datasets. We set the maximum number of possible change points to 5 for the binary segmentation algorithm. Note that this restriction was not applicable to the alternative algorithms provided in the `changepoint` package. In practice, we found that the alternative algorithms tended to over-select the number of change points.

Figure 4(a) and (b) exhibit the change point detection performances for the four scenarios of ℛ_0_. Our BayesSMILES performed much better than the LRT-BinSeg with respect to change point detection under both performance metrics, ARI and MI. For instance, the ARI by BayesSMILES increased 39.29% to 122.16% over the LRT-BinSeg among the four scenarios, while the growth in MI could be up to 60.54%. Figure 4(c) compares the ability to capture the transmission dynamics in terms of RMSE, which depends on the change point detection accuracy. As expected, our BayesSMILES yielded smaller RMSE values across all scenarios since its identified change point locations were more accurate. In all, the simulation study demonstrated the strengths of BayesSMILES.

**Figure 4:**
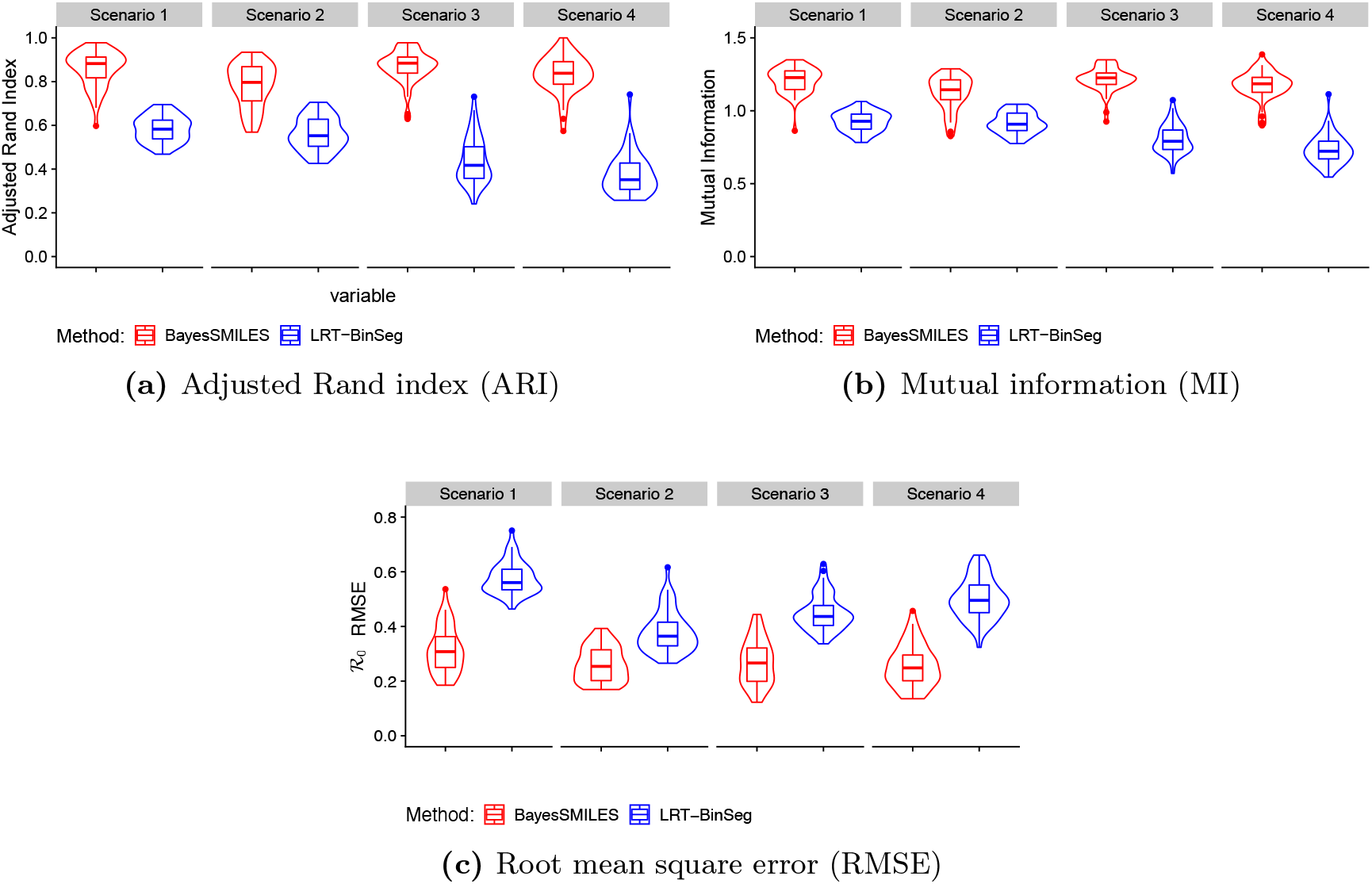
Simulation study: The violin plots of (a) adjusted Rand index, (b) mutual information, and (c) ℛ_0_ root mean square error from 50 replicated datasets generated under the four scenarios. Red and blue violins correspond to the results obtained by BayesSMILES and LRT-BinSeg.

## 6 Analysis of COVID-19 Data

In this section, we applied BayesSMILES to the U.S. state-level COVID-19 daily report data provided by JHU-CSSE COVID-19 Data Repository^1^. Several recent COVID-19 studies also based their analyses on this resource (see e.g. Dong et al., 2020; Zhou and Ji, 2020; Toda, 2020). We first performed a preprocessing step to ensure the quality of the infectious data ***I*** for the model fitting. Due to the fact that recovery cases are not recorded in some states, we treated ***I*** and ***R*** as missing data and reconstructed the two sequences according to the process described in Section 3.1. The cumulative confirmed case numbers ***C*** were collected for each U.S. state starting from an early stage of the pandemic outbreak. In particular, we chose the starting time for each state as when there were at least ten confirmed COVID-19 cases for that state. We also set the removal rate *γ* = 0.1 as suggested by Pedersen and Meneghini (2020) and Weitz et al. (2020). Since different states could have different starting times, we further trimmed the sequences ***I*** and ***R*** for each state based on the latest starting time available. Finally, we set March 22, 2020, as the new starting time (*t* = 1) for all 50 states, and let July 19, 2020, be the last observed time point (*t* = 120).

We used the same hyperparameter and algorithm settings as described in Section 5.3. We ran four MCMC samplers for 40, 000 iterations with the first half discarded as burn-in for the change point detection model to ensure reliable results. We randomly initialized the starting points for each chain. We assessed the concordance between the four chains based on the Pearson correlation coefficients of the marginal posterior probability of inclusions (PPIs), 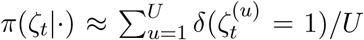. For our real data analysis in this paper, we obtained coefficient values ranging from 0.951 to 0.997, which indicated good concordance among the four MCMC chains. Concordance among the marginal PPIs was confirmed by looking at their scatter plots across each pair of MCMC chains. Furthermore, we also used the Gelman and Rubin’s convergence diagnostics (Gelman et al., 1992) to assess the convergence of the segment-specified basic reproduction numbers ℛ_0*k*_’s to their posterior distributions. The potential scale reduction factors were all below 1.1, ranging from 1.001 to 1.045, clearly indicating that the MCMC chains for the stochastic SIR model were run for a satisfactory number of iterations, which was set to 100, 000. Convergence was also confirmed by looking at their trace plots.

### 6.1 Detecting change points for U.S. states

We limit our analysis to four U.S. states with the highest cumulative confirmed cases as of July 19, 2020, to keep the paper in a reasonable length. They are New York, Texas, California, and Florida. The results for the 46 remaining states are available in https://shuangj00.github.io/BayesSMILES/ (see details in Section 8). Figure 5 displays the detected change points, as well as the estimated basic reproduction number ℛ_0_’s cross segments, for the four states. The associated credible interval to each identified change point is represented by a gray ribbon. In general, those change points detected by BayesSMILES indeed captured the important COVID-19 events that might affect the transmission rates. For instance, some change points reflected the positive effects of the preventative strategies such as lockdown, while others explained the “bounce back” in confirmed cases after reopening. Table 4 lists the change point locations and their potentially related events for the four states.

**Figure 5:**
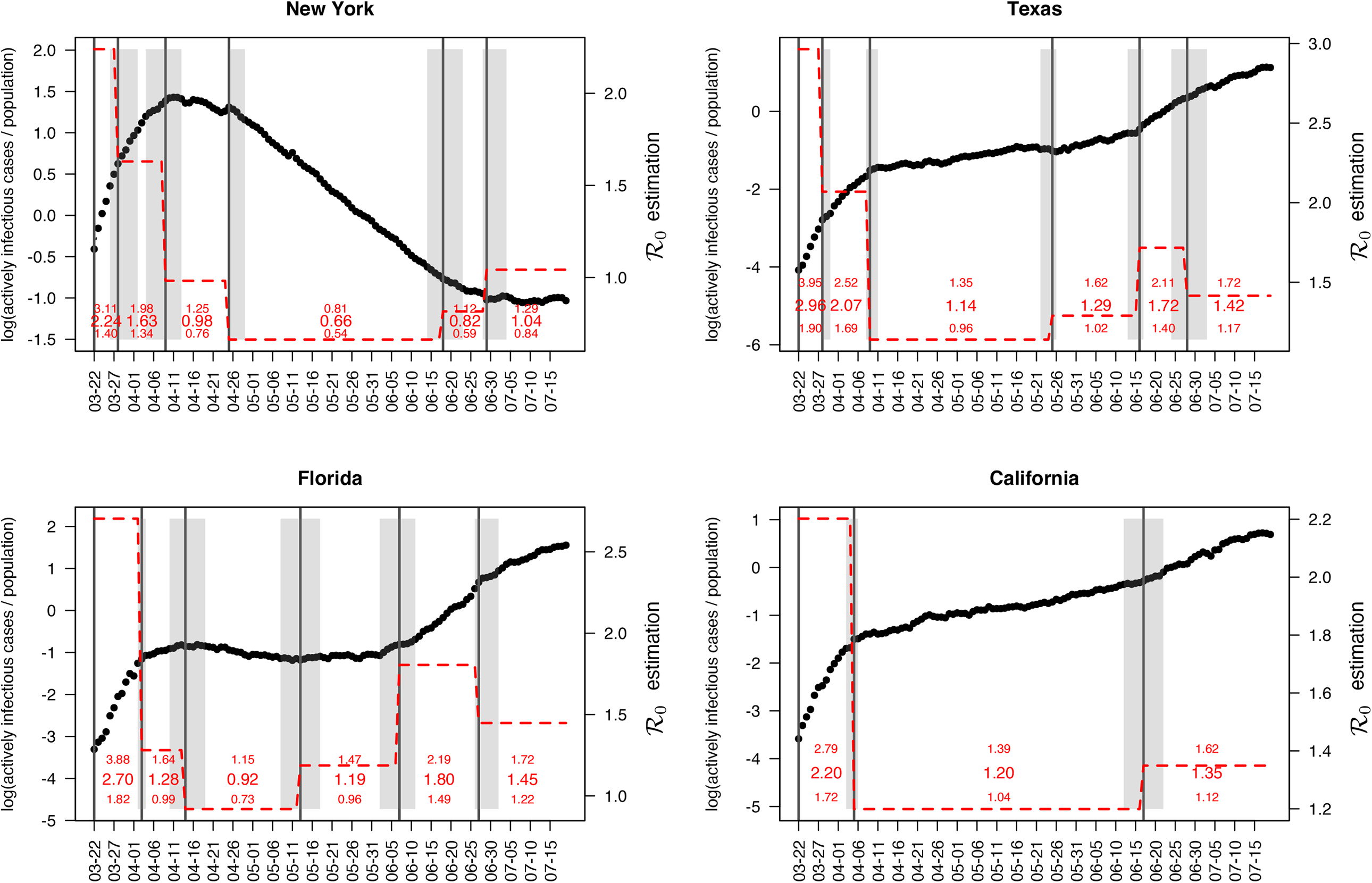
Case study: The change point detection and basic reproduction number estimation for the states of New York, Texas, Florida, and California. The black circles are the actively infectious case numbers divided by the total population (in thousands) at logarithmic scale, i.e. *log*(*I*_*t*_*/N*). The black solid lines pinpoint the change point locations, with the associated gray ribbons indicating the credible intervals. The red dashed lines describe the variation in the basic reproduction numbers ℛ_0_ across segments. The posterior means and the 95% credible intervals for *ℛ*_0*k*_’s are given by red numbers.

In New York, the first change point was estimated to be March 28. We estimated the posterior mean of the basic reproduction number decreased from 2.24 (between March and March 27) to 1.63 (between March 28 and April 8). Notably, March 28 was the date when the Centers for Disease Control and Prevention (CDC) issued a 14-day domestic travel advisory for non-essential persons, which presumably alleviated the situation for the populated states such as New York. The second change point appeared around April 9, and the ℛ_0_ of the third segment dropped to 0.98 with a 95% credible interval of [0.76, 1.25]. This matched the exact day when New York state posted its first drop in the ICU admissions since the COVID-19 outbreak began. The third change point was around April 27. Though there was no direct intervention issued in late April, we noticed that the mayor of New York City announced that all major events had been canceled starting from April 20. This action could bring a positive effect in controlling the outbreak, and our estimation from the SIR model suggested a further decrease in the basic reproduction number down to 0.66 with a 95% credible interval of [0.54, 0.81]. We observed another change point around June 18, which was close to the Phase II reopening of New York state on June 22. During Phase II reopening, restaurants were allowed to open for outdoor dining, stores opened for in-person retail, and more services resumed operational under strict limitations. Thus, we saw a little “bounced back” in ℛ_0_ from 0.66 to 0.82. The last change point was on June 29. As expected, the basic reproduction number increased to 1.04 with a 95% credible interval of [0.84, 1.29] in the last segment. Although there was no public announcement around June 29 with a credible interval from June 28 to July 4, we suspect that the increased social interaction during the Independence Day long weekend (between July 3 and July 5) could be responsible for the increase in transmission dynamics.

In Texas, there were five change points detected. The first change point was estimated to be March 28, the same day as the first one for New York state. Due to a similar reason, the policy of mandatory 14-day quarantines for travelers entering Texas could bring a decrease in terms of the basic reproduction number (decreased from 2.97 to 2.07). The second change point was around April 9 with a further drop of ℛ_0_ to 1.14 with a 95% credible interval [0.96, 1.35]. We found that the Texas Governor had extended the state’s disaster declaration for an additional 30 days on April 12. The extension aimed at protecting the health and safety of Texans by ensuring adequate capabilities of supporting communities. Organizations such as the State Operations Center and the Strategic National Stockpile would continuously supply the state government with the resources needed to protect residents. May 25 was detected as the third change point, and it was the first time that ℛ_0_ increased after the two drops. The estimated basic reproduction number was 1.29 with a 95% credible interval [1.02, 1.62]. This increase appeared around May 25 could be due to the Governor’s updated executive order issued on May 26 that allowed additional services and activities to open for phase II reopening. The next change point was around June 16, and ℛ_0_ further increased to 1.72 with a 95% credible interval [1.40, 2.11]. According to the prediction reported by the University of Texas at Austin’s COVID-19 Modeling Consortium at the end of May, there might be a significant increase in the number of cases and hospitalizations beginning mid-June (News from *kxan*). Here, the change point location and the increased basic reproduction number were consistent with the results of this report. The last change point was around June 28 with an estimated decrease in ℛ_0_ to 1.42 with a 95% credible interval [1.17, 1.72]. Notably, the Texas Governor issued multiple executive orders around late June to early July to mitigate the disease spreading. For instance, the executive order on June 26 reemphasized the limited occupancy for all business establishments in Texas. According to an executive order on July 2, all Texans were required to wear a face-covering in public spaces in counties with 20 or more positive COVID-19 cases. On the same day, the Governor announced an update regarding the executive order on June 26 with additional measures to slow the spread of COVID-19.

In Florida, the first estimated change point was April 3. It was two days after the statewide stay-at-home order for Florida. We estimated that the basic reproduction number decreased from 2.70 to 1.28 after the change point. The second change point appeared around the middle of April. Starting from April 13, some counties such as Osceola county enforced face-covering in public places. It could explain the reason why we observed a slight decrease in ℛ_0_, from 1.28 to 0.92 with a 95% credible interval [0.73, 1.15]. The next change point was located around May 13, and ℛ_0_ in this new stage went above 1 again, with a posterior mean of 1.19 and a 95% credible interval [0.96, 1.50]. We noticed that Florida entered the phase I reopening on May 18, which could lead to the “bounced back” situation. The fourth change point was around June 7, two days after the phase II reopening in Florida. Changes in the phase II reopening included that Universal Orlando opened the parks to the general public for the first time in months, and we observed that ℛ_0_ increased again to 1.81 with a 95% credible interval [1.50, 2.19]. In the last segment (after June 27), our result revealed a slight drop in the basic reproduction number from 1.81 to 1.45. This change was potentially related to the consequence of requiring facial coverings in the four most populated cities in Florida: Tampa, Orlando, Miami, and Jacksonville. The face mask mandates went into effect for the four cities starting from June 19, 20, 25, and 29, respectively. Therefore, the drop in the transmissibility at the end of June may be explained by the effectiveness of wearing face masks as a non-pharmaceutical practice.

In California, we detected two change points. California was the first state to announce lockdown in the COVID-19 pandemic and its stay-at-home order became effective on March 19. Our change point detection results could miss these early actions since the data we analyzed started from March 22. The first selected change point was on April 5, with the value of the basic reproduction number decreasing dramatically when transitioning to the second segment (from 2.20 to 1.20). The second change point was on June 17, and we saw that ℛ_0_ increased to 1.35 in the last segment with a 95% credible interval [1.12, 1.62]. According to California Governor, higher-risk businesses and venues (e.g. movie theaters, bars, gyms) were allowed to reopen with restrictions on June 12. Hence, the increase in the basic reproduction number could be the consequence of reopening. The same observation was made in New York and Texas.

### 6.2 Clustering U.S. states based on their change point locations

We applied BayesSMILES on all 50 U.S. states. Based on the results, we seek to derive an overall picture of the COVID-19 dynamics across states. We summarized the temporally detected change points of the 50 states into common patterns, and then we labeled each state by matching its specific change point pattern to the common patterns. In particular, for each state, we calculated the marginal posterior probability of inclusion (PPI) for all time points, where the PPI for a time point *t* was calculated based on the *B* of MCMC samples after burn-in: 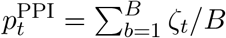. Then we obtained the vector 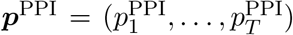. Each entry in ***p***^PPI^ is a value between 0 and 1, representing the proportion of time *t* selected as a change point among all iterations. Next, we computed the overall pattern by averaging over the vector ***p***^PPI^ across 50 states. We noticed that some time points were rarely or never selected as change points. This naturally suggested that we could group the time points. To illustrate this, we trimmed the top 20% values of 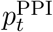 for each time *t* (Figure 6). The trimming step provided a clear pattern and highlighted the groups of dates that were commonly identified as change points. We observed three time spans as shown in Figure 6: March 27 - April 11, May 1 - May 10, and May 22 - July 3. For each state, we defined its cluster label based on the corresponding change point detection results. If a given state had at least one change point (including the credible interval) between March 27 - April 11, the first element in its cluster label is “Change”. Otherwise, the first element in the group label was set to “Stable”. We repeated the same process to determine the second and third elements of the class label for each state. In the end, each state was assigned to a cluster label “Change-Change-Change”, “Change-Stable-Change”, or “Stable-Change-Change”.

**Figure 6:**
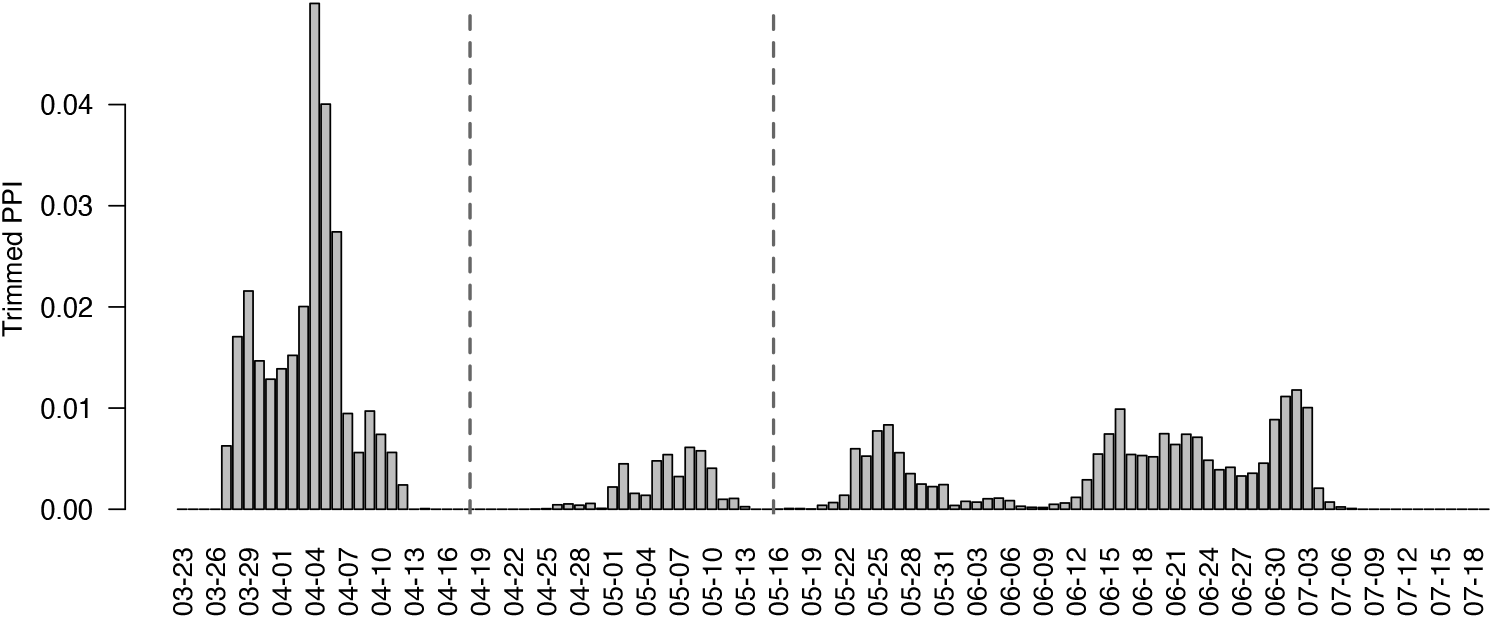
Case study: The averaged marginal posterior probability of inclusion (PPI) for each time point to be selected as a change point over all 50 U.S. states, after trimming the top 20% PPI values. The black dashed lines partition the whole time range into three segments: March 27 - April 11, May 1 - May 10, and May 22 - July 3.

The map in Figure 7 colors each of the 50 states based on its cluster label, where green, yellow, and pink correspond to temporal patterns “Change-Stable-Change”, “Change-Change-Change”, and “Stable-Change-Change”, respectively. Interestingly, three out of the four states we analyzed, New York, Texas, and California, belonged to the same category, “Change-Stable-Change”.Other states also in this category include Georgia, Arizona, North Carolina, and Louisiana. All of these states were in the top ten states with the most COVID-19 confirmed cases. We noticed that the phase I statewide reopening for all these states occurred in mid-May (May 15 for Georgia, May 13 for Arizona, May 8 for North Carolina, May 15 for Louisiana). Therefore, our model did not report any change points for these states between May 1 and May 10. The rest of the 10 states with the most COVID-19 confirmed cases, including Florida, Illinois, and New Jersey, were labeled as “Change-Change-Change”, and all of them had a change point between May 1 and May 10. As discussed in Section 6.1, Florida had a change point around May 13 with a credible interval [May 8, May 18]. According to the Executive Order 2020-32 issued by the Illinois governor, the state entered the phase II reopening starting on May 1 with a modified stay-at-home order. For New Jersey, the statewide state-at-home order was not lifted until June 9. However, our model suggested a change point around the end of April with a credible interval [April 25, May 3] with a drop in ℛ_0_ from 1.11 to 0.68 (details available at https://shuangj00.github.io/BayesSMILES/). We noticed that on May 3 the New Jersey governor announced a multi-state agreement to develop a regional supply chain for personal protective equipment, other medical equipment and testing. This joint-state protective measure allowed for efficient delivery and reliability of medical equipment for states and therefore best utilized life-saving resources in the face of the COVID-19 outbreak.

**Figure 7:**
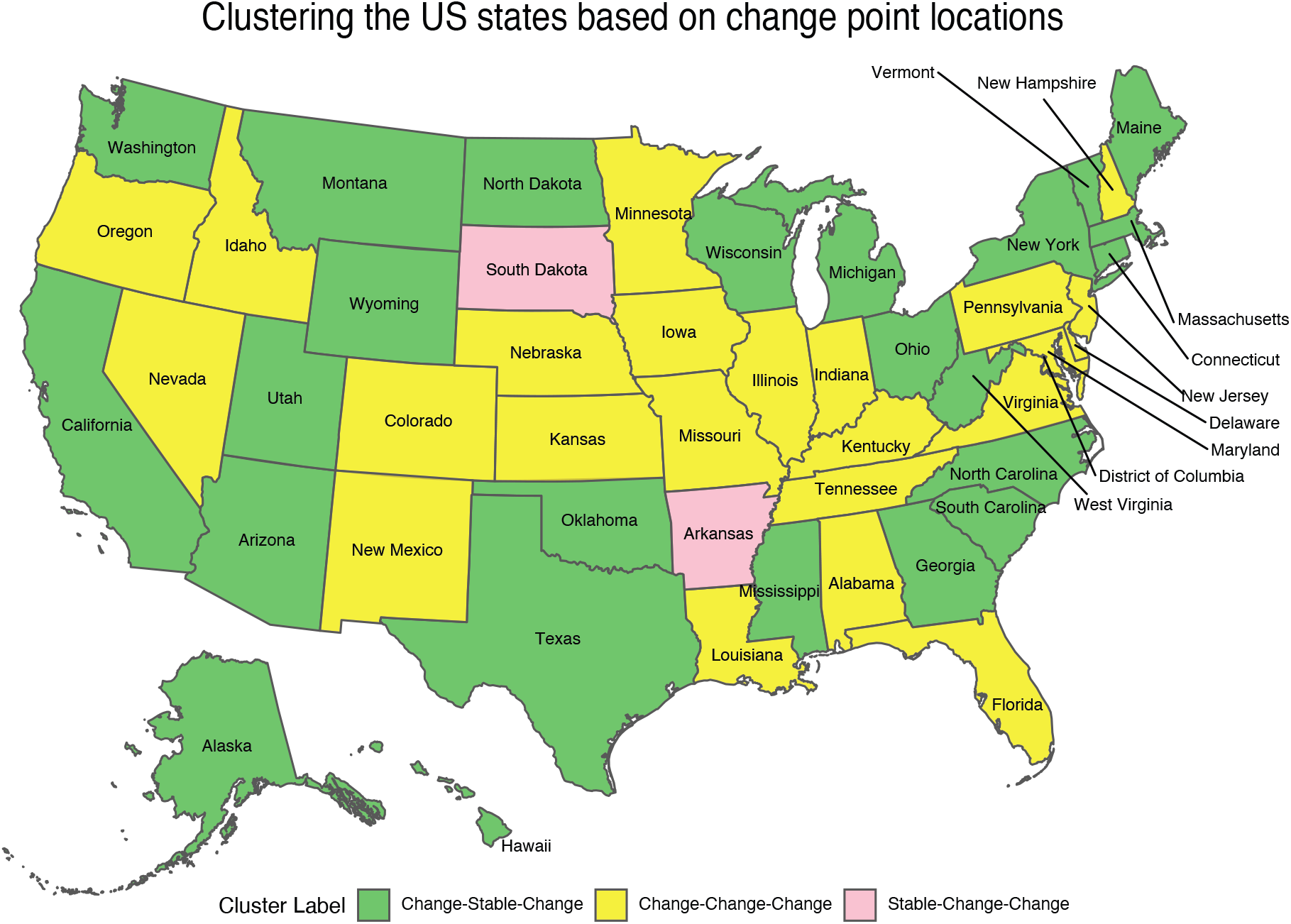
Case study: The temporal patterns of the COVID-19 transmission dynamics based on change points across the 50 U.S. states. Green, yellow, and pink correspond to “Change-Stable-Change”, “Change-Change-Change”, and “Stable-Change-Change” patterns, respectively.

### 6.3 Predicting new confirmed cases for U.S. states

Reliable and accurate short-term forecasting of the new daily confirmed COVID-19 cases 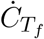 at a future time *T*_*f*_ is important for both policy-makers and healthcare providers. We have illustrated how to use BayesSMILES to predict the new confirmed cases in Section 4.4. The idea is to make the short-term forecast only based only on the observed data in the last available segment, ensuring that only the most recent disease characteristics are utilized. We compared BayesSMILES with the standard stochastic SIR model where all observed data from the first time point are used. We named this model FullDataSIR.

Figure 8 shows the true values of the new daily confirmed COVID-19 cases and the predictions made by BayesSMILES and FullDataSIR for the four major states. The 7-day forecast was chosen from July 20 to 26. First of all, it is observed that the predictive mean by FullDataSIR tended to be larger than that from BayesSMILES. This was because the basic reproduction numbers in the early stage (i.e. from late March to early April) were usually very large due to the lack of effective interventions. As a consequence of including those data, FullDataSIR inflated the predictions. We then quantified the prediction accuracy using the mean absolute percentage error (MAPE). The MAPE for the 7-day forecast is defined as

**Figure 8:**
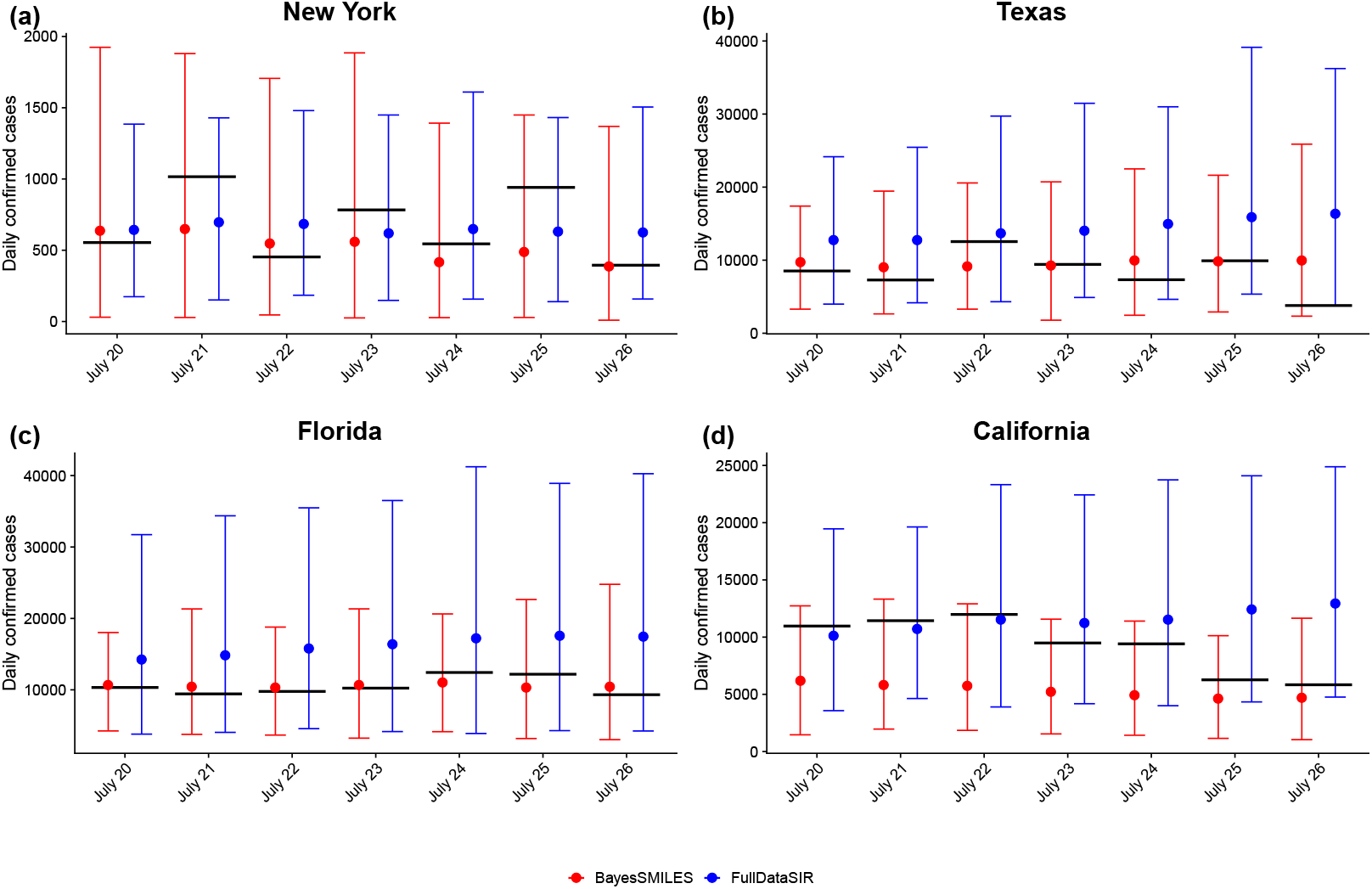
Case study: The 7-day forecast (between July 20 and 26) of the daily confirmed COVID-19 case numbers for the states of New York, Texas, Florida, and California. The red and blue circles and bars are the predictive means and 95% intervals by BayesSMILES and FullDataSIR, respectively. The black thick lines indicate the observed truth.

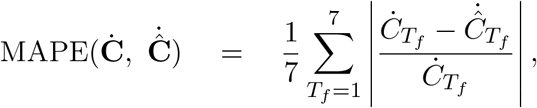

where 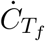and 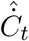 are the observed and predicted new confirmed cases at a future time *T*_*f*_. The smaller the MAPE value, the more accurate the prediction. The numerical summary is shown in Table 3. For New York, Texas, and Florida, the MAPEs from BayesSMILES were much smaller than those from FullDataSIR, suggesting a better performance of BayesSMILES. However, for California, the two methods were almost the same in terms of short-term forecast.

**Table 3:** Case study: The mean absolute percentage errors (MAPEs) of the 7-day forecast of daily confirmed COVID-19 case numbers by BayesSMILES and FullDataSIR for the states of New York, Texas, Florida, and California.

|  | New York | Texas | Florida | California |
| --- | --- | --- | --- | --- |
| BayesSMILES | 24.9 | 38.0 | 8.9 | 40.4 |
| FullDataSIR | 33.0 | 96.7 | 55.2 | 39.9 |

**Table 4:** Case study: The list of the change points identified by BayesSMILES and the related supporting evidences for the states of New York, Texas, Florida, and California.

| State | Change point location<br>(credible interval) | Event | Reference |
| --- | --- | --- | --- |
| New York | Mar 28 (Mar 26 - Apr 2) | 14-day travel advisory | NBC New York (2020a) |
|  | Apr 9 (Apr 4 - Apr 13) | First negative number for ICU admissions | Jesse Pound (2020) |
|  | Apr 27 (Apr 24 - May 1) |  |  |
|  | Jun 18 (Jun 14 - Jun 23) | Phase II reopening | NBC New York (2020b) |
|  | Jun 29 (Jun 28 - Jul 4) |  |  |
| Texas | Mar 28 (Mar 28 - Mar 30) | 14-day travel advisory | Office of the Texas Governor (2020b) |
|  | Apr 9 (Apr 8 - Apr 11) | Texas disaster declaration extension | Ciara Rouege (2020) |
|  | May 25 (May 22 - May 26) | Additional services and activities allowed to open for phase II reopening | Office of the Texas Governor (2020c) |
|  | Jun 16 (Jun 13 - Jun 17) | The second COVID-19 wave in June | Billy Gates (2020) |
|  | Jun 28 (Jun 24 - Jul 3) | Multiple executive orders issued for mitigating the disease spreading | Office of the Texas Governor (2020d)<br>Office of the Texas Governor (2020e)<br>Office of the Texas Governor (2020a) |
| Florida | Apr 3 (Apr 2 - Apr 5) | Statewide stay-at-home order | Klas and Contorno (2020) |
|  | Apr 14 (Apr 10 - Apr 19) | Some counties require face coverings in public | Metevia (2020) |
|  | May 13 (May 8 - May 18) | Phase I reopening | Silcox and Turco (2020) |
|  | Jun 7 (Jun 2 - Jun 11) | Phase II reopening | Dobrzyn (2020) |
|  | Jun 27 (Jun 26 - Jul 2) | Mandatory face coverings in public for major cities | Frago (2020)<br>Speck and Sandoval (2020)<br>The City Manager of the City of Miami (2020)<br>Piggott and McLean (2020) |
| California | Apr 5 (Apr 3 - Apr 6) |  |  |
|  | Jun 17 (Jun 12 - Jun 22) | Higher-risk businesses reopening | Sandhya Kambhampati and Krishnakumar (2020) |

## 7 Conclusion

In this paper, we proposed BayesSMILES, a Bayesian segmentation model for analyzing longitudinal epidemiological data, to characterize the transmission dynamics of an infectious disease such as COVID-19. Our approach includes a Bayesian Poisson segmented regression model to detect multiple change points from the sequence of actively daily infectious cases. Those identified change points correspond to latent events that significantly altered disease spreading rates, while the resulting segments are characterized by unique epidemiological patterns. We further describe the disease transmissibility for each segment by using a stochastic time-invariant SIR model, assuming that the transmission rate remains the same until the next change point. Our model outputs a series of the basic reproduction numbers ℛ_0_’s over stages to track the changes in spreading rates during a pandemic.

We applied BayesSMILES to analyze the COVID-19 daily report data of 50 U.S. states. Our results showed that the COVID-19 outbreak declined substantially after implementing stringent interventions for several states, including New York, Texas, and Florida. Meanwhile, our identified change points matched well with the timelines of publicly announced intervention strategies. The change in the basic reproduction numbers between two adjacent segments might be used to quantify the effectiveness of an intervention, which could help us understand the impact of different control measures. Several downstream analyses based on the BayesSMILES results were conducted. In particular, we clustered the temporal patterns of the 50 U.S. states based on their change point locations, which led to an interesting spatial pattern related to the COVID-19 dynamics. Lastly, we demonstrated that our method could also improve the short-term forecasting of the new daily confirmed cases.

A potential issue of BayesSMILES is that the change point locations, which are identified in the Poisson segmented regression model, are set to be fixed when estimating the basic reproduction numbers using the stochastic SIR model. Such a two-stage approach might under-estimate the uncertainties in ℛ_0*k*_’ s. A diagnostic method named simulation-based calibration (SBC) (Talts et al., 2018) is available to assess if the model inference has properly quantified the uncertainty. Using SBC to evaluate the soundness of the current MCMC sampling methods could be a future exploration. Another potential extension of the current work is to utilize advanced versions of the MH algorithm in the MCMC algorithms. For example, the MH with delayed rejection (Mira et al., 2001), the combination of delayed rejection and adaptive Metropolis samplers (Haario et al., 2006), the multiple-try Metropolis (Liu et al., 2000; Martino, 2018), as well as the methods discussed in Liang et al. (2011).One may also extend the Poisson error structure in the change point detection model to a negative binomial distribution for modeling the over-dispersed count data. Furthermore, the current BayesSMILES framework can be generalized to characterize temporal patterns in other epidemiological data. To do so, the segmented regression model should not be restricted to countable outcomes. Due to the concern for data accuracy, the result provided by the proposed method must be interpreted with caution. For instance, the number of confirmed cases is largely dependent on the test capacity and the number of recovery cases may suffer from under-reporting issues. How to improve the statistical power and prediction accuracy under those circumstances is worth investigating.

## 8 Software

We provide software in the form of `R/C++` codes on GitHub https://github.com/shuangj00/BayesSMILES. It includes the tutorial of implementing BayesSMILES, using U.S. state-level COVID-19 data as an example. Besides, we have designed a website https://shuangj00.github.io/BayesSMILES/ to summarize the inference results for the 50 U.S. states, as a supplement to Section 6. The website shows that 1) the detected change points for each U.S. state; and 2) the COVID-19 transmission dynamics based on the segment-varying basic reproduction numbers ℛ_0_’s, including their posterior means and 95% credible intervals.

## Data Availability

The COVID-19 dataset was downloaded from the COVID-19 Data Repository hosted by Johns Hopkins University Center for Systems Science and Engineering

https://github.com/CSSEGISandData/COVID-19

## 9 Acknowledgment

This work was supported by the University of Texas at Dallas (UT Dallas) Office of Research [UT Dallas Center for Disease Dynamics and Statistics] and partially supported by the National Institutes of Health [5P30CA142543, 5R01GM126479, 5R01HG008983]. The authors would like to thank Jessie Norris for proofreading the manuscript.

## Appendix

### A1. Approximate the multivariate normal density function

This section provides the details of the multivariate normal density approximation used to improve the computational efficiency in Section 4.1. We consider a general setting as follows. Let ***y*** be an *n* × 1 vector, ***W*** be an *n* × *q* matrix, ***U*** be an *n* × *p* matrix, and ***X*** = (***W***, ***U***) (which is an *n* × (*p* + *q*) matrix). Let **Σ** be a (*q* + *p*) × (*q* + *p*) diagonal matrix where the first *q* diagonal elements are *h*_0_ and the last *p* diagonal elements are *h*_1_. Suppose *h*_0_, *h*_1_, *σ*^2^ *>* 0. By Woodbury identity and Sylvester’s determinant identity, we have

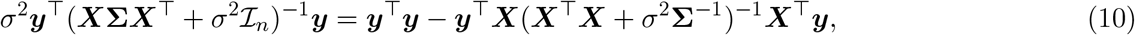

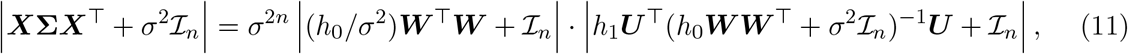

where | · | denotes the matrix determinant and ℐ_*n*_ is an n-dimensional diagonal matrix. Define

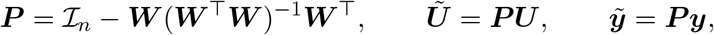

where ***Ũ*** (respectively 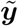) is the residual after regressing out ***W*** from ***U*** (respectively ***y***). Zhou and Guan (2018) showed that the expressions in (10) and (11) can be further simplified when *h*_0_ →∞. The results are summarized below with the proof available in the supplement of Zhou and Guan (2018).

#### Lemma 1.

*Let* 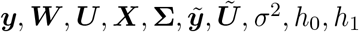, *be defined as above. Then*,

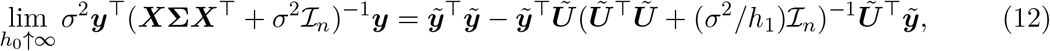

*and*

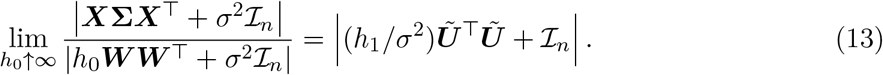

The conclusions in Lemma 1 can be used to improve the computational efficiency for approximating the multivariate normal probability density function, shown in Equation (7), in our model. Within each segment *k* we assumed 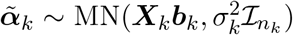. Under the prior specification discussed in Section 3.3, we have 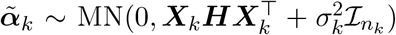, and the corresponding p.d.f. for 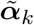 is:

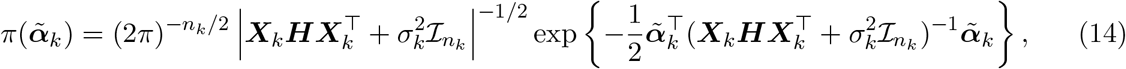

where *n*_*k*_ is the segment length. Next, we simplify the calculation of 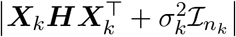 and 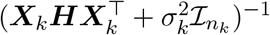 by using Lemma 1. Consider 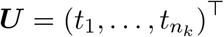 and let ***W*** be a column vector of 1’s with length *n*_*k*_. The vector ***y*** in our case matches 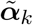 and 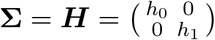. Lemma 1 states that the inverse and determinant calculation of an *n*_*k*_ × *n*_*k*_ matrix can be reduced to that of a *p* × *p* matrix, and in our case *p* = 1 (since the regression model only includes “time” as a covariate except the intercept term). Therefore, the computational benefit could be significant when *n*_*k*_ was large. We first derive the formula of ***P*** as follows,

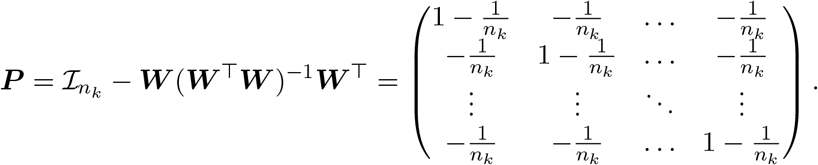

Next, according to Equation (12), 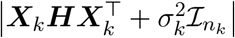 can be approximated by 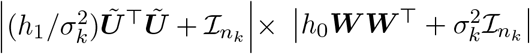 when *h*_0_ → +∞. In particular, we can derive

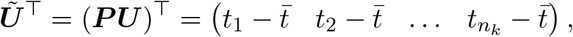

where 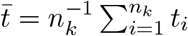 denotes the average. Therefore,

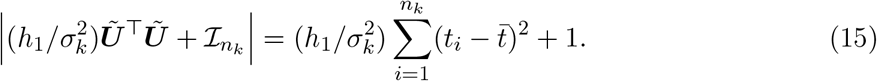

To calculate 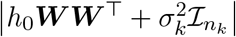, upon noting that ***WW*** ^*T*^ is an *n*_*k*_ × *n*_*k*_ matrix, we find

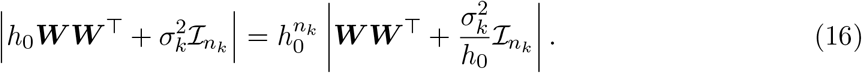

According to Sylvester’s determinant lemma, we have

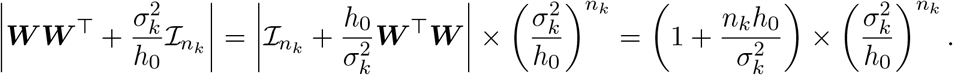

Therefore, the formula in Equation (16) equals the following,

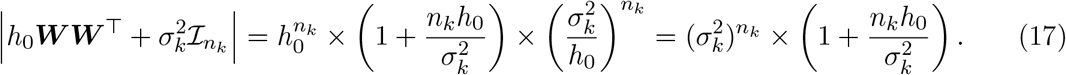

Combining the results in Equations (15) and (17), we can approximate the matrix determinant in (14) as follows when *h*_0_ → ∞,

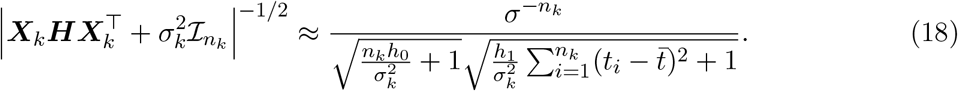

Next, it is straightforward to derive the formula in the exponent part in Equation (14) using the result in Equation (12). In particular, we can derive:

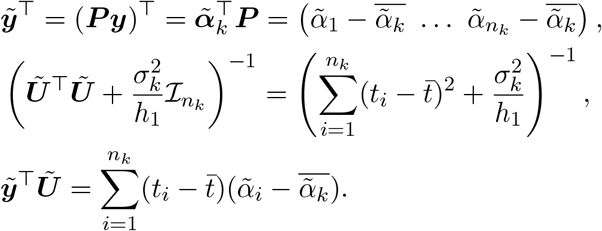

Then we approximate the exponent part in Equation (14) as follows under the condition that *h* → ∞,

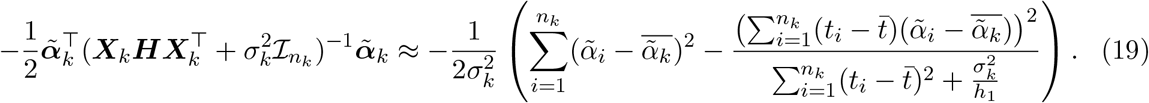

We further introduce the following notation,

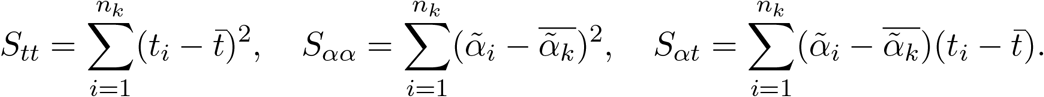

By combining the approximations in (18) and (19), we obtain

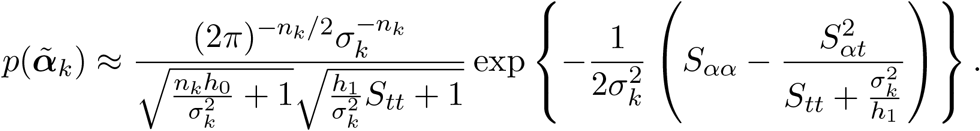

In practice, assuming that all covariates are standardized, this approximation works well if *h*_0_*/h*_1_ is large enough. We suggest using *h*_0_ = 10, 000 and *h*_1_ = 10, which yielded highly accurate approximations in our simulation.

### A2. Results for Single Simulated Dataset

This section provides additional simulation results for Scenarios 1, 2, and 3 described in Section 5.1. For each scenario, we randomly selected a single simulated dataset from the 50 replicates. Figure 9 shows the PPM estimates on the change point indicator ***ζ***. The red dashed and blue solid lines represent the true and estimated change point locations, respectively, while the gray ribbons indicate the corresponding 95% credible intervals. As we can see, BayesSMILES correctly pointed out the true change points. The resulting ARI values were 0.87, 0.85, and 0.95, respectively, while the MI values were 1.21, 1.18, and 1.31, respectively. Figure 10 shows our estimates on the basic reproduction number ℛ_0*k*_ for each segment partitioned by the identified change points. The red dashed and blue solid line pinpoint the true and posterior mean of 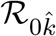’s, while the two black solid lines mark the boundary of 95% credible intervals. In all scenarios, the true values were within their corresponding 95% credible intervals. The ℛ_0_ RMSEs for the single datasets from the three scenarios were 0.33, 0.36, and 0.14, respectively.

**Figure 9:**
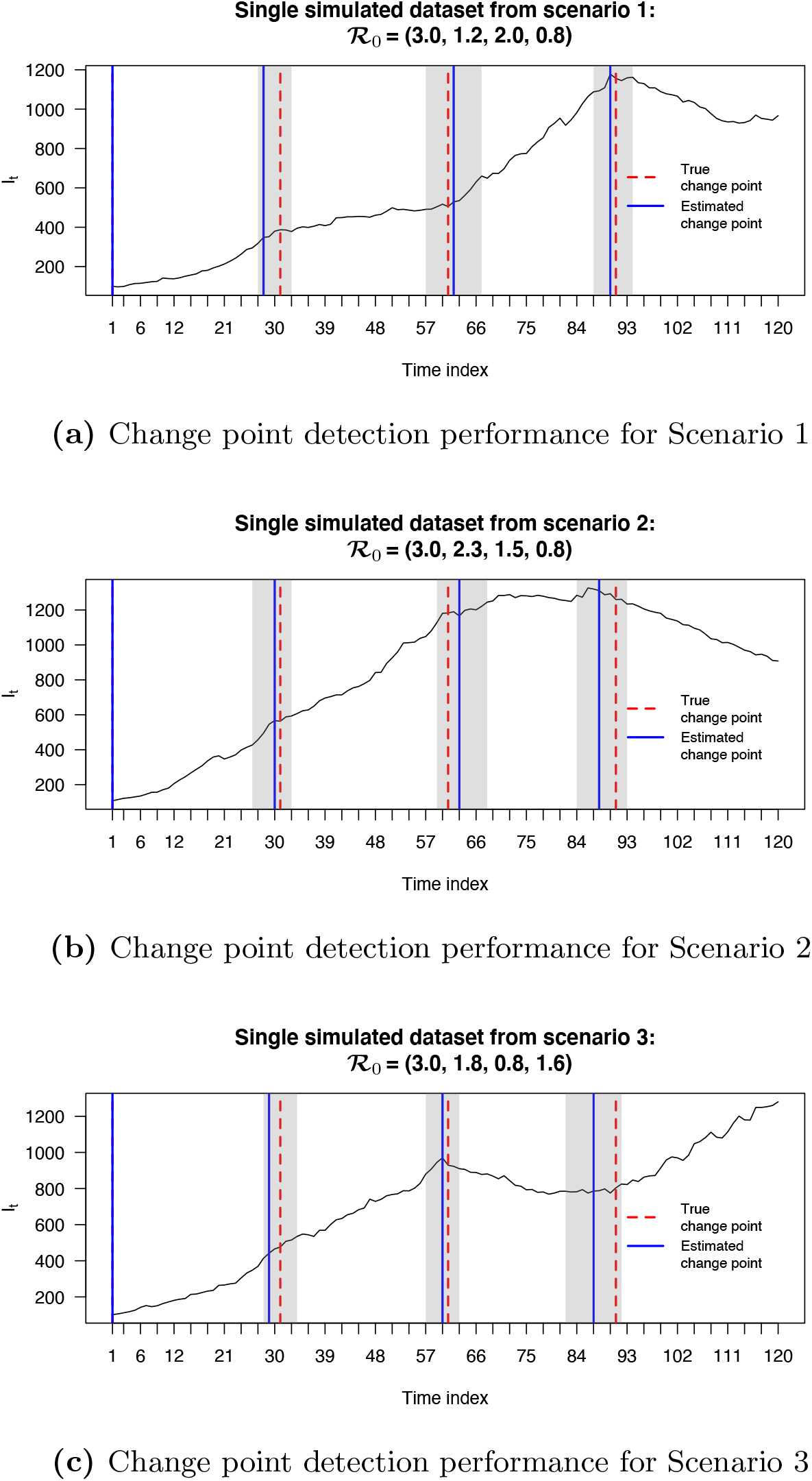
Simulation study: The locations of change points (blue solid lines) estimated from the posterior pairwise probability matrix (PPM) and their credible intervals (gray ribbons). The red dashed lines mark the true change point locations.

**Figure 10:**
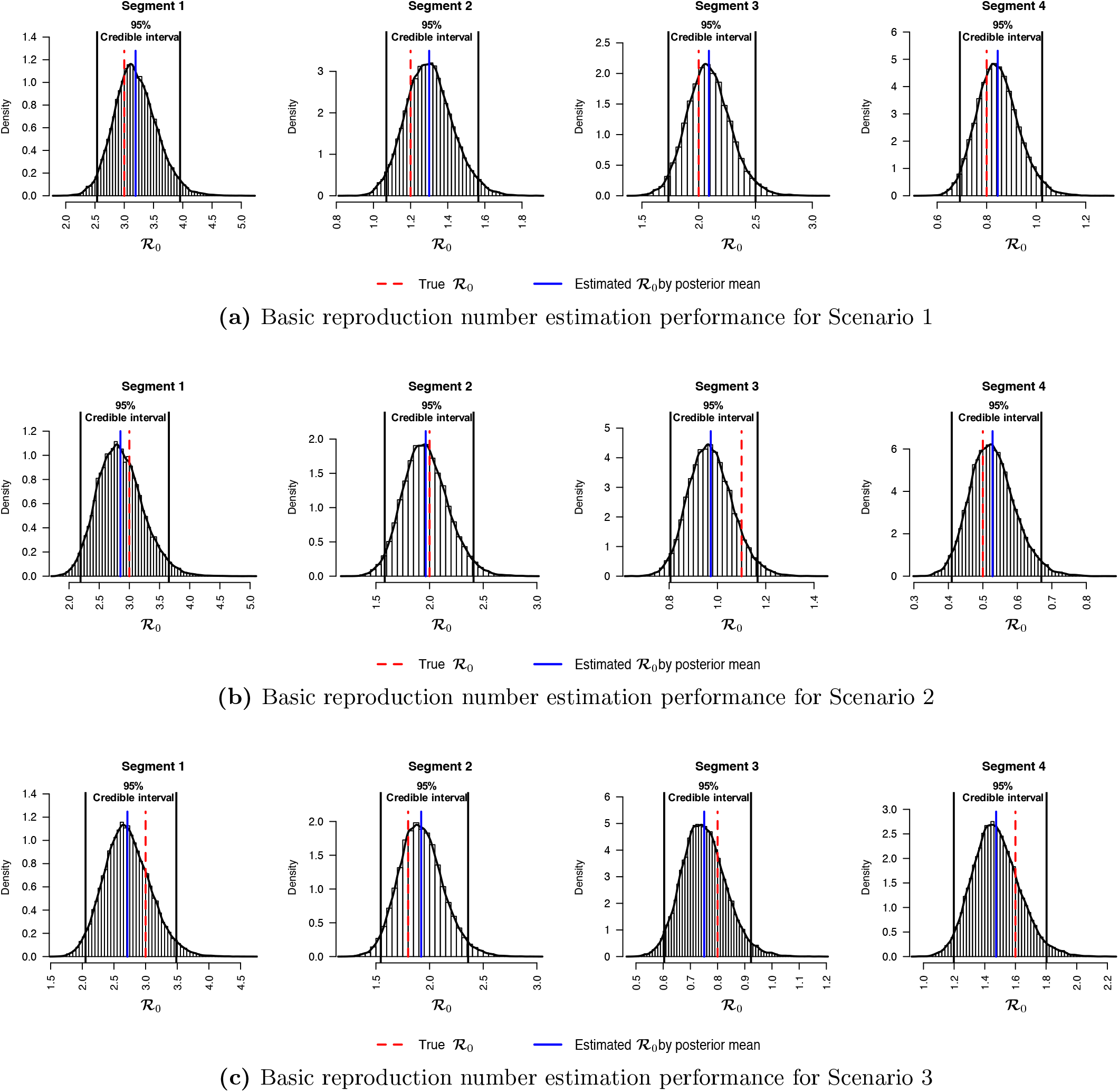
Simulation study: The posterior distributions of ℛ_0*k*_’s for *k* = 1, 2, 3, 4 estimated from the segmented time-series data, given the three identified change points as shown in Figure 9. The red dashed and blue solid lines are the true and estimated values of ℛ_0*k*_’s, respectively. The two black solid lines are the lower and upper bounds of the 95% credible intervals.

## Footnotes

1 https://github.com/CSSEGISandData/COVID-19

## References

Akiyama MJ, Spaulding AC, Rich JD (2020). Flattening the curve for incarcerated populations—COVID-19 in jails and prisons. New England Journal of Medicine, 382(22): 2075–2077.

Allen LJ (2008). An introduction to stochastic epidemic models. In: Mathematical Epidemiology, 81–130. Springer.

Alvarez FE, Argente D, Lippi F (2020). A simple planning problem for COVID-19 lockdown. Technical report, National Bureau of Economic Research.

Andersson H, Britton T (2012). Stochastic epidemic models and their statistical analysis, volume 151. Springer Science & Business Media.

Bailey NT, et al. (1975). The mathematical theory of infectious diseases and its applications.

Charles Griffin & Company Ltd, 5a Crendon Street, High Wycombe, Bucks HP13 6LE. Becker NG, Britton T (1999). Statistical studies of infectious disease incidence. Journal of the Royal Statistical Society: Series B (Statistical Methodology), 61(2): 287–307.

Billy Gates AGYC Matthew Prendergast (2020). Austin health leaders concerned about possible 2nd COVID-19 wave in June. kxan. URL: https://www.kxan.com/news/austin-public-health-to-give-covid-19-update-at-9-a-m/, accessed 2020-05-27.

Chen YC, Lu PE, Chang CS (2020). A Time-dependent SIR model for COVID-19. arXiv preprint arXiv:2003.00122.

Chowell G, Castillo-Chavez C, Fenimore PW, Kribs-Zaleta CM, Arriola L, Hyman JM (2004). Model parameters and outbreak control for SARS. Emerging Infectious Diseases, 10(7): 1258.

Ciara Rouege (2020). Abbott extends Texas disaster declaration | Here’s what that means. KHOU. URL: https://www.khou.com/article/news/health/coronavirus/gov-abbott-extends-disaster-declaration-another-30-days/285-9e494ee3-5ea5-48d9-bf35-b8c4b88f5126, accessed 2020-04-12.

Cooper I, Mondal A, Antonopoulos CG (2020). A SIR model assumption for the spread of COVID-19 in different communities. Chaos, Solitons & Fractals, 139: 110057.

Dehning J, Zierenberg J, Spitzner FP, Wibral M, Neto JP, Wilczek M, et al. (2020). Inferring change points in the spread of COVID-19 reveals the effectiveness of interventions. Science.

Dobrzyn E (2020). Gov. Ron DeSantis says most of Florida will enter phase 2 of reopening Friday. Click Orlando. URL: https://www.clickorlando.com/news/local/2020/06/03/gov-ron-desantis-says-most-of-florida-will-enter-phase-2-of-reopening-friday/, accessed 2020-06-03.

Dong E, Du H, Gardner L (2020). An interactive web-based dashboard to track COVID-19 in real time. The Lancet Infectious Diseases, 20(5): 533–534.

Edwards AW, Cavalli-Sforza LL (1965). A method for cluster analysis. Biometrics, 362–375.

Flaxman S, Mishra S, Gandy A, Unwin HJT, Mellan TA, Coupland H, et al. (2020). Estimating the effects of non-pharmaceutical interventions on COVID-19 in Europe. Nature, 584(7820): 257–261.

Frago C (2020). Tampa’s mask order went into effect Friday. Here’s what you need to know. Tampa Bay News. URL: https://www.tampabay.com/news/tampa/2020/06/19/tampas-mask-order-goes-into-effect-today-heres-what-you-need-to-know/, accessed 2020-06-19.

Gelman A, Rubin DB, et al. (1992). Inference from iterative simulation using multiple sequences. Statistical Science, 7(4): 457–472.

Giordano G, Blanchini F, Bruno R, Colaneri P, Di Filippo A, Di Matteo A, et al. (2020). Modelling the COVID-19 epidemic and implementation of population-wide interventions in Italy. Nature Medicine, 1–6.

Gostic KM, McGough L, Baskerville E, Abbott S, Joshi K, Tedijanto C, et al. (2020). Practical considerations for measuring the effective reproductive number,Rt. medRxiv.

Haario H, Laine M, Mira A, Saksman E (2006). DRAM: efficient adaptive MCMC. Statistics and Computing, 16(4): 339–354.

Hinkley DV (1970). Inference about the change-point in a sequence of random variables.

Hubert L, Arabie P (1985). Comparing partitions. Journal of Classification, 2(1): 193–218.

Jen T, Gupta AK (1987). On testing homogeneity of variances for Gaussian models. Journal of Statistical Computation and Simulation, 27(2): 155–173.

Jesse Pound (2020). New York posts negative net change in ICU admissions for first time since coronavirus outbreak. CNBC. URL: https://www.cnbc.com/2020/04/10/ny-has-first-negative-net-change-in-icu-admissions-since-coronavirus-outbreak.html, accessed 2020-04-10.

Kantner M, Koprucki T (2020). Beyond just “flattening the curve”: Optimal control of epidemics with purely non-pharmaceutical interventions. Journal of Mathematics in Industry, 10(1): 1–23.

Kermack WO, McKendrick AG (1927). A contribution to the mathematical theory of epidemics. Proceedings of the royal society of london. Series A, Containing papers of a mathematical and physical character, 115(772): 700–721.

Killick R, Eckley I (2014). changepoint: An R package for changepoint analysis. Journal of Statistical Software, 58(3): 1–19.

Killick R, Fearnhead P, Eckley IA (2012). Optimal detection of changepoints with a linear computational cost. Journal of the American Statistical Association, 107(500): 1590–1598.

Klas ME, Contorno S (2020). Florida Gov. Ron DeSantis issues statewide stay-at-home order. Tampa Bay News. URL: https://www.tampabay.com/news/health/2020/04/01/florida-gov-ron-desantis-issues-statewide-stay-at-home-order/, accessed 2020-04-

Liang F, Liu C, Carroll R (2011). Advanced Markov chain Monte Carlo methods: learning from past samples, volume 714. John Wiley & Sons.

Liu JS, Liang F, Wong WH (2000). The multiple-try method and local optimization in Metropolis sampling. Journal of the American Statistical Association, 95(449): 121–134.

Lloyd-Smith JO, Galvani AP, Getz WM (2003). Curtailing transmission of severe acute respiratory syndrome within a community and its hospital. Proceedings of the Royal Society of London. Series B: Biological Sciences, 270(1528): 1979–1989.

Martino L (2018). A review of multiple try MCMC algorithms for signal processing. Digital Signal Processing, 75: 134–152.

Metevia T (2020). Osceola County now requires face coverings in public. Click Orlando. URL: https://www.clickorlando.com/news/local/2020/04/10/watch-live-osceola-county-officials-provide-update-on-covid-19-pandemic/, accessed 2020-04-13.

Mira A, et al. (2001). On Metropolis-Hastings algorithms with delayed rejection. Metron, 59(3-4): 231–241.

NBC New York (2020a). CDC Issues 14-Day Travel Advisory for New York, New Jersey, Connecticut. NBC New York. URL: https://www.nbcnewyork.com/news/local/even-with-relief-bill-passed-no-rest-for-ny-as-cuomo-says-peak-of-crisis-still-yet-to-come/2348306/, accessed 2020-03-28.

NBC New York (2020b). NYC Moves to Phase II in Biggest Reopening Step Yet; State Daily Virus Deaths Plunge to New Low. NBC New York. URL: https://www.nbcnewyork.com/news/local/nyc-reopening-hits-highest-gear-yet-as-shops-outdoor-dining-and-playgrounds-return-monday/2477627/, accessed 2020-06-22.

Office of the Texas Governor (2020a). A proclamation amending Executive Order GA-28 by Office of the Texas Governor. URL: https://open.texas.gov/uploads/files/organization/opentexas/DISASTER-proclamation-amending-GA-28-mass-gatherings-IMAGE-07-02-2020.pdf, accessed 2020-07-02.

Office of the Texas Governor (2020b). Executive Order GA-11 by Office of the Texas Governor. URL: https://www.nbcnewyork.com/news/local/nyc-reopening-hits-highest-gear-yet-as-shops-outdoor-dining-and-playgrounds-return-monday/2477627/, accessed 2020-03-26.

Office of the Texas Governor (2020c). Executive Order GA-24 by Office of the Texas Governor. URL: https://gov.texas.gov/uploads/files/press/DISASTER_Adding_Covered_Services_to_GA-23_No_2_COVID-19.pdf, accessed 2020-05-26.

Office of the Texas Governor (2020d). Executive Order GA-28 by Office of the Texas Governor. URL: https://gov.texas.gov/uploads/files/press/EO-GA-28_targeted_response_to_reopening_COVID-19.pdf, accessed 2020-06-26.

Office of the Texas Governor (2020e). Executive Order GA-29 by Office of the Texas Governor. URL: https://open.texas.gov/uploads/files/organization/opentexas/EO-GA-29-use-of-face-coverings-during-COVID-19-IMAGE-07-02-2020.pdf, accessed 2020-07-02.

Pedersen MG, Meneghini M (2020). Quantifying undetected COVID-19 cases and effects of containment measures in italy. ResearchGate Preprint (online 21 March 2020) DOI, 10.

Piggott J, McLean J (2020). Jacksonville changes course, issues face mask mandate. News4JAX. URL: https://www.news4jax.com/news/local/2020/06/29/jacksonville-issues-face-mask-mandate/, accessed 2020-06-29.

Rand WM (1971). Objective criteria for the evaluation of clustering methods. Journal of the American Statistical Association, 66(336): 846–850.

Riley S, Fraser C, Donnelly CA, Ghani AC, Abu-Raddad LJ, Hedley AJ, et al. (2003). Transmission dynamics of the etiological agent of SARS in Hong Kong: impact of public health interventions. Science, 300(5627): 1961–1966.

Riou J, Hauser A, Counotte MJ, Althaus CL (2020). Adjusted age-specific case fatality ratio during the COVID-19 epidemic in Hubei, China, January and February 2020. MedRxiv, 2020.

Sandhya Kambhampati MM, Krishnakumar P (2020). Which California counties are reopening? Los Angeles Times. URL: https://www.latimes.com/projects/california-coronavirus-cases-tracking-outbreak/reopening-across-counties/.

Santos JM, Embrechts M (2009). On the use of the adjusted rand index as a metric for evaluating supervised classification. In: International Conference on Artificial Neural Networks, 175–184. Springer.

Sen A, Srivastava MS (1975). On tests for detecting change in mean. The Annals of Statistics, 98–108.

Silcox F, Turco R (2020). Florida Businesses Ready for Full Phase One Reopening. Bay News. URL: https://www.baynews9.com/fl/tampa/coronavirus/2020/05/18/ongoing-coronavirus-coverage, accessed 2020-05-18.

Song PX, Wang L, Zhou Y, He J, Zhu B, Wang F, et al. (2020). An epidemiological forecast model and software assessing interventions on COVID-19 epidemic in China. medRxiv.

Speck E, Sandoval E (2020). Orange County residents will be required to wear face masks under new executive order. Click Orlando. URL: https://www.clickorlando.com/news/local/2020/06/18/orange-county-residents-will-be-required-to-wear-face-masks-under-new-executive-order/, accessed 2020-06-18.

Steuer R, Kurths J, Daub CO, Weise J, Selbig J (2002). The mutual information: detecting and evaluating dependencies between variables. Bioinformatics, 18(suppl_2): S231–S240.

Tadesse MG, Sha N, Vannucci M (2005). Bayesian variable selection in clustering highdimensional data. Journal of the American Statistical Association, 100(470): 602–617.

Talts S, Betancourt M, Simpson D, Vehtari A, Gelman A (2018). Validating bayesian inference algorithms with simulation-based calibration. arXiv preprint arXiv:1804.06788.

The City Manager of the City of Miami (2020). City Mandates Facial Coverings in Public; Civil Penalties Approved. URL: https://www.miamigov.com/files/sharedassets/public/news/2020/0625-emergency-order-20-16.pdf, accessed 2020-06-25.

Toda AA (2020). Susceptible-infected-recovered (SIR) Dynamics of COVID-19 and Economic Impact. arXiv preprint arXiv:2003.11221.

Vehtari A, Gelman A, Gabry J (2017). Practical Bayesian model evaluation using leave-one-out cross-validation and WAIC. Statistics and Computing, 27(5): 1413–1432.

Waqas M, Farooq M, Ahmad R, Ahmad A (2020). Analysis and Prediction of COVID-19 Pandemic in Pakistan using Time-dependent SIR Model. arXiv preprint arXiv:2005.02353.

Weitz JS, Beckett SJ, Coenen AR, Demory D, Dominguez-Mirazo M, Dushoff J, et al. (2020). Modeling shield immunity to reduce COVID-19 epidemic spread. Nature Medicine, 1–6.

Zhou Q, Guan Y (2018). On the null distribution of bayes factors in linear regression. Journal of the American Statistical Association, 113(523): 1362–1371.

Zhou T, Ji Y (2020). Semiparametric Bayesian Inference for the Transmission Dynamics of COVID-19 with a State-Space Model. arXiv preprint arXiv:2006.05581.

